# Uncertainty extraction and verification for the Common Conditions Affecting the Hand and Wrist Priority Setting Partnership: A proposed methodology for Priority Setting Partnerships with the James Lind Alliance

**DOI:** 10.1101/2021.03.18.21253314

**Authors:** DJC Grindlay, TRC Davis, D Kennedy, D Larson, D Furniss, K Cowan, G Giddins, A Jain, RW Trickett, A Karantana

## Abstract

**Objective:** To report our recommended methodology for extracting and confirming published research uncertainties during a broad scope Priority Setting Partnership (PSP) with the James Lind Alliance.

**Setting:** This process was completed in the UK as part of the PSP for “Common Conditions Affecting the Hand and Wrist”, comprising of health professionals, patients and carers.

**Design and Data Extraction:** We applied best practice in systematic review methodology and reporting, including independent screening and data extraction by multiple researchers and use of a PRISMA flowchart, alongside steering group consensus processes.

**Data Sources:** Cochrane Systematic Reviews and Protocols, NICE (National Institute for Health and Care Excellence) Guidelines, and SIGN (Scottish Intercollegiate Guidelines Network) Guidelines.

**Selection Criteria:** Selection of research uncertainties was guided by the scope of the Common Conditions Affecting the Hand and Wrist PSP which focused on “common hand conditions routinely treated by hand specialists, including hand surgeons and hand therapists limited to identifying questions concerning the results of intervention, and not the basic science or epidemiology behind disease.

**Results:** Of 2358 records identified after duplicate extraction which entered the screening process, 186 records were presented to the PSP steering group for eligibility assessment; 79 were deemed within scope and included for the purpose of research uncertainty extraction (45 full Cochrane Reviews, 18 Cochrane Review protocols, 16 Guidelines). These yielded 89 research uncertainties, which were compared to the stakeholder survey, and added to the longlist where necessary; before derived uncertainties were checked against non-Cochrane published systematic reviews.

**Conclusions:** In carrying out this work, beyond reporting on output of the Common Conditions Affecting the Hand and Wrist PSP, we detail the methodology and processes we hope can inform and facilitate the work of future PSPs and other evidence reviews, especially those with a broader scope beyond a single disease or condition.

**Strengths and Limitations:**

– This is a novel and rigorous approach to uncertainty extraction and verification in the setting of a James Lind Alliance Priority Setting Partnership.
– The detailed methodology provides transparency regarding the extraction and verification of uncertainties.
– The uncertainty extraction and verification relied upon the HandSRev database – similar resources may not be available for other Priority Setting Partnerships.

## BACKGROUND

The James Lind Alliance (JLA) ^1^ is a non-profit making initiative that brings together patients, carers and frontline clinicians to identify and prioritise research uncertainties through a Priority Setting Partnership (PSP) ^2^. A PSP highlights to funding bodies those research topics which are most important to patients and clinicians alike.

In 2017, The Common Conditions Affecting the Hand and Wrist PSP, funded by the British Society for Surgery of the Hand (BSSH) and managed by a multi-centre, multidisciplinary steering group, reported the top prioritised research uncertainties to map out a framework for future clinical research in common hand conditions. The broader process and results of this PSP are published separately^3^.

The current article focuses on principles guiding the identification and verification of uncertainties from the published literature. These were considered in the Partnership long list alongside the uncertainties directly submitted through the stakeholder survey. Collating these published uncertainties was especially challenging given the huge scope of hand surgery, stretching across both plastic and orthopaedic surgery parent specialties and a wide range of conditions and injuries.

Whilst the methods of collating stakeholder uncertainties (either via surveys or interviews with patients, carers, and clinicians) are well defined, a reproducible methodology for extracting uncertainties from the published literature, and for the verification of submitted uncertainties, has not yet been proposed by the JLA^4^.

As the JLA PSP methodology becomes widely adopted, an increasing number of partnerships focus on broader research areas ^5-9^. A lack of framework to enable accurate and efficient extraction of research uncertainties is a hurdle for such broad scope PSPs.

## Objective

This study reports the processes of identifying potential research uncertainties in the published literature in guidelines and Cochrane Reviews, and verifying uncertainties for the Common Conditions Affecting the Hand and Wrist Priority Setting Partnership. In doing so, it aims to formalise a robust and reproducible method of identifying and confirming research priority uncertainties in such a setting.

## METHODS

The full PSP methodology for collating submitted research questions is detailed in the full PSP output and executive report ^3^.

### Patient and Public Involvement

The PSP Steering Group (SG) included five patient representatives with personal experience of treatment for common hand surgery conditions, who helped shape the process throughout, with emphasis on defining the scope and informing the consensus.

### Selection criteria

Selection of research uncertainties for the prioritisation process was guided by the scope of the Common Conditions Affecting the Hand and Wrist PSP, which is detailed in the final report ^3^. In summary the scope focused on “common hand conditions that are routinely treated by hand specialists, including hand surgeons and hand therapists” ^3^. In line with the established JLA methods at the time, the selection was limited to identifying research questions concerning the results of intervention, and not the basic science or epidemiology behind disease ^4^.

### Sources and search strategy

The PSP Steering Group (SG) specified that existing uncertainties in the published literature would be identified via the following sources:

- Cochrane Systematic Reviews
- Cochrane Systematic Review Protocols
- NICE (National Institute for Health and Care Excellence) Guidelines and
- SIGN (Scottish Intercollegiate Guidelines Network) Guidelines.

Cochrane Systematic Reviews and Protocols were chosen because of the consistent, high quality of their methodology. These were identified by searching the Cochrane Library (http://www.cochranelibrary.com/), using a comprehensive search strategy for hand surgery conditions and terms developed by DG for the Centre for Evidence Based Hand Surgery at the University of Nottingham (Supplementary file 1). Where a Cochrane Review had been updated, the most recent version was used. Withdrawn reviews and withdrawn review protocols were excluded.

NICE and SIGN national guidelines relevant to the scope of the PSP were identified using the NHS Evidence search engine (https://www.evidence.nhs.uk/), again using a comprehensive search strategy relevant to hand surgery conditions and terms. The limitation to the length of the search strategy that could be used in NHS Evidence meant that the search had to be split into four separate searches that were run separately. For each search the filter for “Guidance” was applied, combined successively with “National Institute for Health and Care Excellence – NICE” and “Scottish Intercollegiate Guidelines Network – SIGN” from the “Source” option. In the case of guidelines on the same topic generated by both NICE and SIGN, each was considered independently for uncertainly extraction. Withdrawn guidelines were excluded.

The search date was 12 September 2016. There were no date restrictions applied. Results were combined and duplicates removed.

### Selection process

Two members of the SG (AK/DG) independently screened search results to identify all guidelines and Cochrane Reviews potentially relevant to conditions affecting the hand and wrist. The identified references were collated and grouped into broad categories and presented to the SG for screening and ratification. The SG ratified, via consensus, references as relevant to the scope of the PSP, which pertains to “common conditions affecting the hand and wrist”. Where disagreement existed between steering group members, this was resolved via a two-stage consensus process. The first stage consisted of each of eleven non-lay SG members independently screening identified relevant records for eligibility. The second involved discussion between all SG members for contentious reports in a face-to-face meeting. Consensus for each stage was achieved when percentage agreement between SG members was 75% or more ^10^. The process was documented via a PRISMA (Preferred Reporting Items for Systematic Reviews and Meta-Analyses) flow chart ^11^.

### Research uncertainty extraction

The Cochrane Reviews, Cochrane Review protocols and NICE and SIGN guidelines determined to be within scope of the PSP by the above process were then distributed amongst the clinical members of the SG for extraction of research uncertainties. Members worked in pairs. The process was piloted, using two example reviews and two reviewers. Reviewers were tasked with forming a research uncertainty from any research recommendations or demonstrable gaps in knowledge highlighted in the identified documents.

In identifying uncertainties from Cochrane Reviews, emphasis was placed on the “Authors’ conclusions”, including sections on “Implications for practice” and “Implications for research”. To avoid over-interpretation of the data and encourage transparency, reviewers were asked to focus, wherever possible, on explicit statements of treatment uncertainties and research needs.

Each identified uncertainty was accompanied by an explicit statement(s), extracted verbatim from the text, of whether or not there was particular evidence for a research question (review) and/or if the uncertainty was identified by a guideline group as a research priority (guidelines). Examples of such statements are:

> *“Research is required to determine the duration of benefit from local corticosteroid injection and to identify candidates for treatment based on severity and duration of symptoms*.*”*

> *“Local corticosteroid injection should also be compared to, and combined with, other non-surgical and even surgical interventions to determine the optimum management of carpal tunnel syndrome*.*”*

Reviewers were advised to look out for phrases such as “there is insufficient evidence”, “the evidence is insufficient”, “there is no evidence that”, etc., as possible clues to uncertainties. Care was taken to distinguish cases where a series of good quality RCTs had shown little or no evidence of the efficacy of a treatment (which implies a certainty of lack of effectiveness rather than an uncertainty about effectiveness) from cases where reliable RCTs were lacking (which implies an uncertainty).

Each pair of reviewers identified potential areas of uncertainty independently before discussing their choices as a pair. Once agreement was achieved, a final uncertainty was submitted to the SG. If agreement was not reached, then all versions of the uncertainty were submitted and ratified separately by the SG. In compiling uncertainties, the PICO model was considered whenever applicable, to construct clinical questions: Patient, Population or Problem; Intervention; Comparison; Outcome ^12^.

Where apparent from the systematic review or guidelines, elements of PICO such as specific patient groups or comparator treatments were built into the uncertainties. For example, a systematic review may have found no evidence concerning effectiveness of a treatment in a specific group of patients or in a particular condition. In these cases, specific uncertainties to match these PICO criteria would be appropriate.

For ease, in the title field for each uncertainty, the Intervention (I) was mentioned first, followed by the Population/Patient (IP); for example, Surgery (I) in adults with carpal tunnel syndrome (P). If the uncertainty included comparators and/or specified outcomes, the Intervention was mentioned first, followed by the Comparator (C), followed by the Population/Patient (ICP); for example, surgery (I) compared with splints (C) in adults with carpal tunnel syndrome (P). If outcomes were specified in the uncertainty, these were included [ICPO].

Uncertainties extracted by each pair of reviewers were recorded in a table proforma, compatible with the general format of the standard JLA data management Excel spreadsheet ^13^. Generated uncertainties were assembled, categorised and duplicates removed (RT/AK/DG).

### Research uncertainty verification

Research uncertainty verification was performed by the non-lay members of the SG, working in pairs.

The aim of verification was not to repeat the work of Cochrane reviewers or guideline developers by re-assessing the quality of individual studies included in each record. However, to increase transparency, SG members were asked to review the results section and make a judgement as to why each uncertainty constituted a persistent research gap by selecting one of five options (A-E) as detailed in Table 1.

**Table 1.**
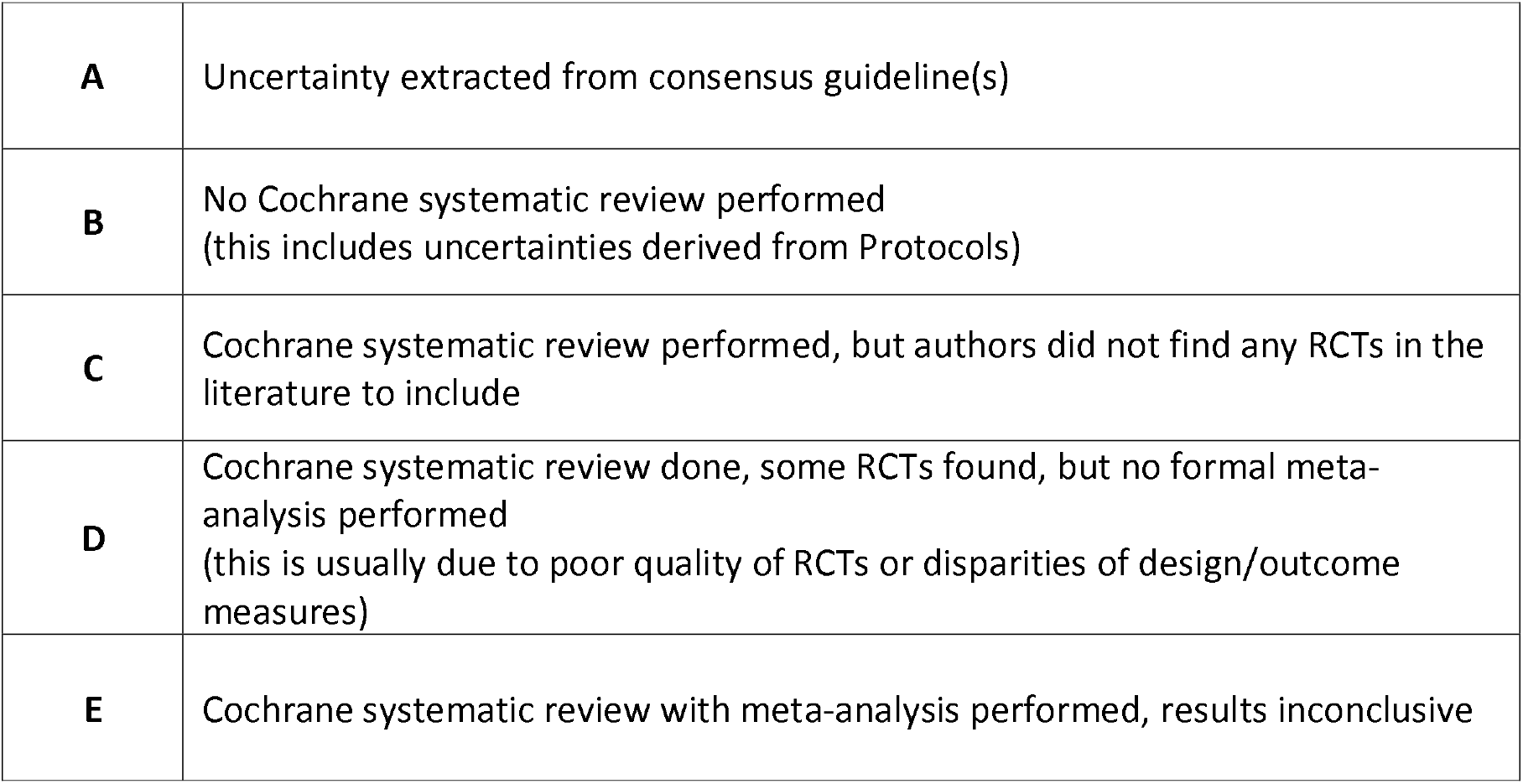
Categories used to document the level of uncertainty at source

|  |  |
| --- | --- |
| **A** | Uncertainty extracted from consensus guideline(s) |
| **B** | No Cochrane systematic review performed<br>(this includes uncertainties derived from Protocols) |
| **C** | Cochrane systematic review performed, but authors did not find any RCTs in the literature to include |
| **D** | Cochrane systematic review done, some RCTs found, but no formal meta-analysis performed<br>(this is usually due to poor quality of RCTs or disparities of design/outcome measures) |
| **E** | Cochrane systematic review with meta-analysis performed, results inconclusive |

As most of the included Cochrane Reviews and guidelines were potentially out of date (more than 3 years from publication), each identified uncertainty was further checked against the preceding five years (2011 −2016) of published non-Cochrane systematic reviews, as identified through the HandSRev database ^14^. HandSRev is an open-access database of systematic reviews relevant to hand surgery practice, mapped by clinical themes, compiled and updated by the Centre for Evidence Based Hand Surgery (CEBHS) at the University of Nottingham. New systematic reviews are identified monthly using a comprehensive search strategy developed by DG and AK for PubMed.

The final list of uncertainties derived from the published literature was ratified by the SG.

## RESULTS

### Report selection

Results of the screening and two-stage consensus process are illustrated in the PRISMA flow chart in Figure 1 ^11^. A total of 2358 records identified through the searches entered the screening process after duplicate extraction. After first screening and exclusion of records that were obviously out of scope, 186 records were presented to the full SG for eligibility assessment as described in the methods. Seventy nine records were deemed within scope of the PSP by this process and were included for the purpose of research uncertainty extraction. These consisted of 45 full Cochrane Reviews, 18 Cochrane Review protocols and 16 Guidelines and are listed in Supplementary file 2, grouped by source and clinical topic.

**1.**
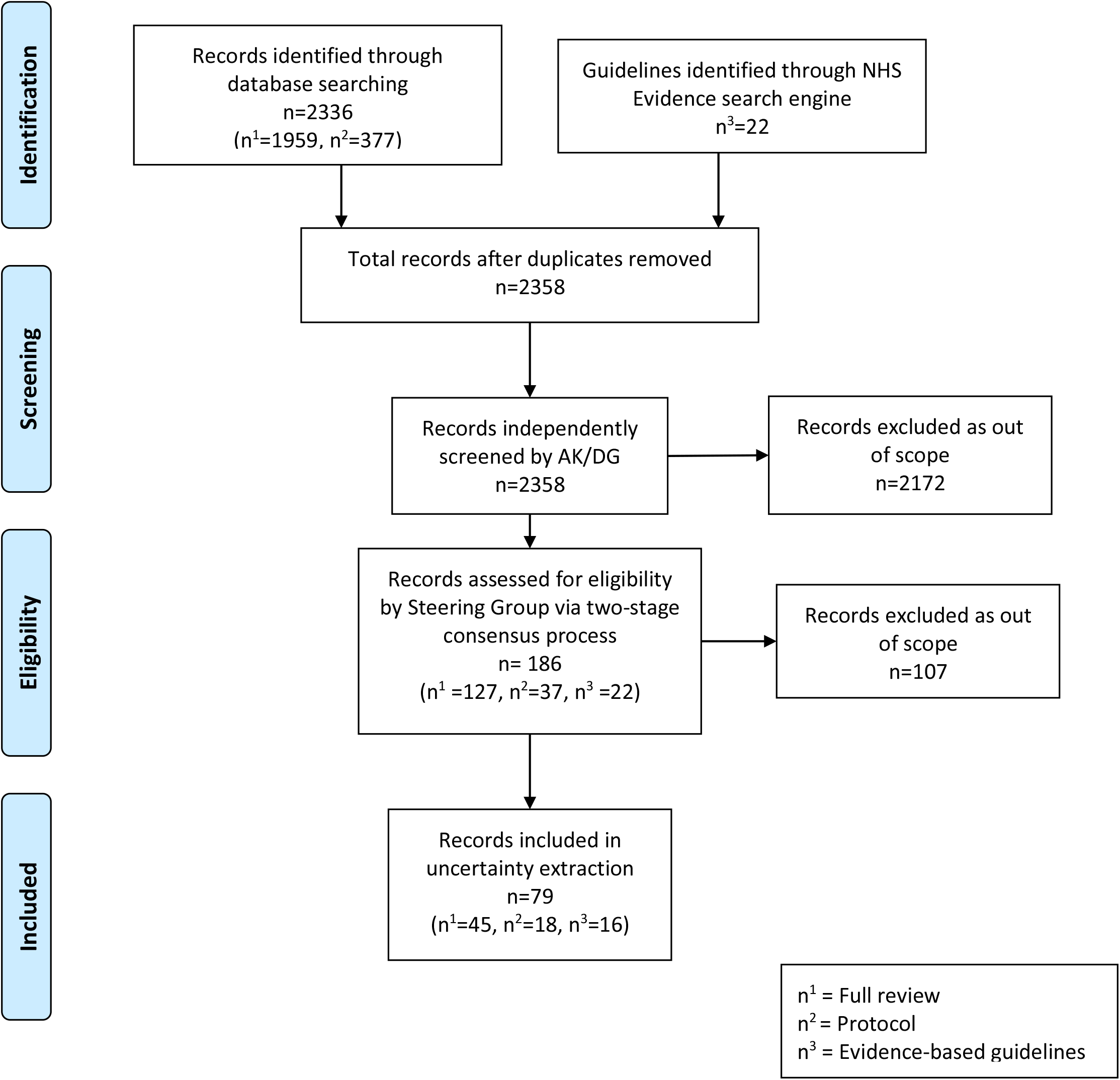
PRISMA flow chart of uncertainty extraction

### Uncertainly extraction and verification

The results of the uncertainty extraction process for this PSP are presented in Supplemental file 3. This details the published source, the level of uncertainly (as per Table 1), the parameters of the PICO, the original verbatim uncertainty text as extracted from the source and the derived uncertainty in PICO format wherever possible. The 79 records yielded 89 research uncertainties. These were cross referenced with uncertainties submitted via the stakeholder survey and further checked against the relevant output from the HandsRev database any evidence updates since their date of publication. Finally they were ratified by the SG for inclusion into the PSP longlist (Supplemental file 4) ^15^.

## DISCUSSION

We applied best practice in systematic review methodology and reporting, including independent screening and data extraction by multiple researchers and use of a PRISMA flowchart, alongside steering group consensus processes, with the aim to maximise transparency and reduce potential for bias.

Until recently, most JLA PSPs have focused on a single disease (e.g. stroke, eczema) - see https://www.jla.nihr.ac.uk/priority-setting-partnerships. At the time of set up, the Common Conditions Affecting the Hand and Wrist PSP was unusual in that it was aimed at the entirety of a medical specialty area (hand surgery and hand therapy in general), making its breadth extremely broad. The scope included a very wide range of conditions and intervention types. This made the identification and extraction of uncertainties from the relevant published resources in a systematic and reproducible way challenging.

While not all PSPs extract uncertainties from published literature and guidelines, all are required to verify that questions submitted via surveys are indeed true uncertainties. This verification process requires the findings from up-to-date systematic reviews, where available. It is hoped that the process described here will help others do this in a more structured, systematic and reproducible way. With such a wide range of conditions and procedures to cover, it is key to have comprehensive search strategies that will find all relevant evidence. The PSP was able to draw on the exhaustive PubMed search strategy developed by the Centre for Evidence Based Hand Surgery for the HandSRev database of systematic reviews relevant to hand surgery practice ^14^. This strategy has several hundred terms and includes hand disease and hand surgery terms, combined with a systematic review filter. Ideally for any PSP a similar comprehensive search strategy covering the full scope is required to identify all the relevant systematic reviews.

As Cochrane reviews and evidence-based guidelines require methodological rigour and thus take a long time to produce, an unintended consequence is that they can become superseded as new evidence appears. In this work, the HandSRev database ^14^, was also used to check that the final questions submitted to the PSP longlist were up-to date genuine research uncertainties, as it is updated monthly with the results of new searches. A number of speciality specific databases for other conditions, are available online. However, the setup and upkeep of such resources is resource intensive and requires funds. Hence, for many cross-cutting PSPs, it is unlikely there will be a specialty-specific database available to easily identify relevant systematic reviews. In such cases it would be necessary to develop a comprehensive search strategy from scratch, and then compile a list of systematic reviews by topic.

An potential alternative source for PSPs identifying and verifying uncertainties has emerged with the advent of the Epistemonikos database (https://www.epistemonikos.org/), which is a free online database of systematic reviews. Epistemonikos is multilingual database of health evidence which regularly screens multiple electronic databases and other resources, resulting in the largest source of systematic reviews relevant for health-decision making, maintained by a non-profit organisation founded in 2009 ^16^. However, it would still be necessary to compile and report a comprehensive search strategy, with all relevant diseases and terms relevant to the PSP’s remit, for Epistemonikos and any other resources used (e.g. NICE Evidence Search). Hence input of an experienced information specialist remains key to the evidence review and uncertainty verification processes of a PSP.

In carrying out this work, beyond reporting on the details of the output of the Common Conditions Affecting the Hand and Wrist PSP ^3^, we developed and detailed the methodology and processes which we hope can inform and facilitate the work of future PSPs and/or other evidence reviews, especially those with a broader scope beyond a single disease or condition.

## Data Availability

Raw data available at http://www.jla.nihr.ac.uk/priority-setting-partnerships/common-conditons-affecting-the-hand-and-wrist/

## AKNOWLEDGEMENTS

All steering group patient members: Christopher Delaney, Jo Horsey, David Skilton, Sheila Wade, and Gillian Walton, and those clinicians and patients who volunteered their valuable time for submission of questions, responding to surveys, or participating in the final workshop. The British Society for Surgery of the Hand for funding the PSP.

## Funding declaration

This Priority Setting Partnership was funded in full by the British Society for Surgery of the Hand (BSSH).

## Conflict of Interest

This work was funded by the British Society for Surgery of the Hand.

AK, AJ, GG, DF, TRCD, and RWT are consultant hand surgeons and members of the British Society for Surgery of the Hand. All are active in clinical and non-clinical research.

No authors received compensation or renumeration in anyway for the work completed.

KC is a senior adviser to the James Lind Alliance and co-author and editor of the JLA Guidebook. She supports the JLA Secretariat including in recruiting and training the team of JLA advisers. She was an independently contracted adviser for this work.

None of the authors declare any other competing interests.

## Patient and Public Involvement

The entirety of this work involved patients throughout, in line with the James Lind Alliance objectives.

## Contributor Statement

Origin of study idea – DG, RWT, AK. Acquisition of data – DG, AK. Analysis of data – RWT. Interpretation of data – AK, TRCD, DK, DL, DF, DG, KC, GG, AJ, RWT. Drafting and revision – AK, TRCD, DK, DL, DF, DG, KC, GG, AJ, RWT. Final approval – AK, TRCD, DK, DL, DF, DG, KC, GG, AJ, RWT. Agreement of accountability – AK, TRCD, DK, DL, DF, DG, KC, GG, AJ, RWT.

## Notes

### Author Declarations

no relevant ethical guidelines to follow, this was a JLA PSP

### Summary of Updates

changed corresponding author email address to institutional email rather than personal

