## Supplemental file 1 for "Uncertainty extraction and verification for the Common Conditions Affecting the Hand and Wrist Priority Setting Partnership: A proposed methodology for Priority Setting Partnerships with the James Lind Alliance"

**Supplemental file 1: JLA Cochrane Library search strategy**

Search date: 12th September 2016

hand:ti,ab,kw or hands:ti,ab,kw or wrist or wrists or finger or fingers or fingertip or fingertips or thumb or thumbs or volar or elbow or elbows or forearm or forearms or "upper limb" or carpal or metacarpal or phalanx or phalanges or phalangeal or interphalangeal or trapeziometacarpal or metacarpophalangeal or cubital or scaphoid or radius or radial or ulna or ulnar or "extensor tendon" or "flexor tendon" or "triangular fibrocartilage" or colles or colles's or mallet or dupuytren or dupuytren's or epicondylitis or tenosynovitis or tendinopathy or tendinitis or trigger or quervain or quervain's or pulley or club or clubbed or ganglion or "brachial plexus" or "complex regional pain" or CRPS or causalgia or [mh hand] or [mh "hand joints"] or [mh "hand bones"] or [mh wrist] or [mh "wrist joint"] or [mh metacarpus] or [mh fingers] or [mh "finger joint"] or [mh thumb] or [mh forearm] or [mh "capitate bone"] or [mh "carpal bones"] or [mh "hamate bone"] or [mh "lunate bone"] or [mh "metacarpal bones"] or [mh "finger phalanges"] or [mh "pisiform bone"] or [mh "radius"] or [mh "scaphoid bone"] or [mh "trapezium bone"] or [mh "trapezoid bone"] or [mh "triquetrum bone"] or [mh ulna] or [mh "olecranon process"] or [mh "triangular fibrocartilage"] or [mh "brachial plexus"] or [mh "hand transplantation"] or [mh "nails"] or [mh "hand deformities"] or [mh "polydactyly"] or [mh "syndactyly"] or [mh "brachydactyly"] or [mh "de quervain disease"] or [mh "trigger finger disorder"] or [mh "tenosynovitis"] or [mh "ganglion cysts"] or [mh "dupuytren contracture"] or [mh "carpal tunnel syndrome"] or [mh "cubital tunnel syndrome"] or [mh "tennis elbow"] or [mh "hand injuries"] or [mh "finger injuries"] or [mh "radius fractures"] or [mh "ulna fractures"] or [mh "colles' fracture"] or [mh "monteggia's fracture"] or [mh "paronychia"] or [mh "complex regional pain syndromes"]

Note: Protocols have no MeSH terms added
Note: By default search includes full text
