## Supplemental file 2 for "Uncertainty extraction and verification for the Common Conditions Affecting the Hand and Wrist Priority Setting Partnership: A proposed methodology for Priority Setting Partnerships with the James Lind Alliance"

**Supplementary file 2: Included records, deemed within scope and submitted for research uncertainty extraction**

**Cochrane Reviews**

*n = 45*

**Carpal tunnel syndrome**

Surgical treatment options for carpal tunnel syndrome.
Cochrane Database of Systematic Reviews, no. 4 (2007)
http://onlinelibrary.wiley.com/doi/10.1002/14651858.CD003905.pub3/abstract

Surgical versus non-surgical treatment for carpal tunnel syndrome.
Cochrane Database of Systematic Reviews, no. 4 (2008)
http://onlinelibrary.wiley.com/doi/10.1002/14651858.CD001552.pub2/abstract

Endoscopic release for carpal tunnel syndrome.
Cochrane Database of Systematic Reviews, no. 1 (2014)
http://onlinelibrary.wiley.com/doi/10.1002/14651858.CD008265.pub2/abstract

Splinting for carpal tunnel syndrome.
Cochrane Database of Systematic Reviews, no. 7 (2012)
http://onlinelibrary.wiley.com/doi/10.1002/14651858.CD010003/abstract

Local corticosteroid injection for carpal tunnel syndrome.
Cochrane Database of Systematic Reviews, no. 2 (2007)
http://onlinelibrary.wiley.com/doi/10.1002/14651858.CD001554.pub2/abstract

Non-surgical treatment (other than steroid injection) for carpal tunnel syndrome.
Cochrane Database of Systematic Reviews, no. 1 (2003)
http://onlinelibrary.wiley.com/doi/10.1002/14651858.CD003219/abstract

Therapeutic ultrasound for carpal tunnel syndrome.
Cochrane Database of Systematic Reviews, no. 3 (2013)
http://onlinelibrary.wiley.com/doi/10.1002/14651858.CD009601.pub2/abstract

Rehabilitation following carpal tunnel release.
Cochrane Database of Systematic Reviews, no. 2 (2016)
http://onlinelibrary.wiley.com/doi/10.1002/14651858.CD004158.pub3/abstract

Exercise and mobilisation interventions for carpal tunnel syndrome.
Cochrane Database of Systematic Reviews, no. 6 (2012)
http://onlinelibrary.wiley.com/doi/10.1002/14651858.CD009899/abstract

Ergonomic positioning or equipment for treating carpal tunnel syndrome.

Cochrane Database of Systematic Reviews, no. 1 (2012)
http://onlinelibrary.wiley.com/doi/10.1002/14651858.CD009600/abstract

**Cubital tunnel syndrome & ulnar neuropathy/nerve compression**

Treatment for ulnar neuropathy at the elbow.
Cochrane Database of Systematic Reviews, no. 11 (2016)
http://onlinelibrary.wiley.com/doi/10.1002/14651858.CD006839.pub3/abstract

**Compression neuropathies other than carpal/cubital tunnel**

Treatment for thoracic outlet syndrome.
Cochrane Database of Systematic Reviews, no. 11 (2014)
http://onlinelibrary.wiley.com/doi/10.1002/14651858.CD007218.pub3/abstract

**Stroke & adult spasticity**

Hands-on therapy interventions for upper limb motor dysfunction following stroke.
Cochrane Database of Systematic Reviews, no. 6 (2011)
http://onlinelibrary.wiley.com/doi/10.1002/14651858.CD006609.pub2/abstract

Interventions for improving upper limb function after stroke.
Cochrane Database of Systematic Reviews, no. 11 (2014)
http://onlinelibrary.wiley.com/doi/10.1002/14651858.CD010820.pub2/abstract

**Cerebral palsy**

Surgical treatment for the thumb-in-palm deformity in patients with cerebral palsy.
Cochrane Database of Systematic Reviews, no. 4 (2005)
http://onlinelibrary.wiley.com/doi/10.1002/14651858.CD004093.pub2/abstract

**De Quervain's disease**

Corticosteroid injection for de Quervain's tenosynovitis.
Cochrane Database of Systematic Reviews, no. 3 (2009)
http://onlinelibrary.wiley.com/doi/10.1002/14651858.CD005616.pub2/abstract

**Dupuytren's contracture**

Surgery for Dupuytren's contracture of the fingers.
Cochrane Database of Systematic Reviews, no. 12 (2015)

http://onlinelibrary.wiley.com/doi/10.1002/14651858.CD010143.pub2/abstract

**Trigger digits**

Corticosteroid injection for trigger finger in adults.
Cochrane Database of Systematic Reviews, no. 1 (2009)
http://onlinelibrary.wiley.com/doi/10.1002/14651858.CD005617.pub2/abstract

**Osteoarthritis of the hand**

Surgery for thumb (trapeziometacarpal joint) osteoarthritis.
Cochrane Database of Systematic Reviews, no. 2 (2015)
http://onlinelibrary.wiley.com/doi/10.1002/14651858.CD004631.pub4/abstract

**Inflammatory conditions and arthropathies**

*Rheumatoid arthritis & wrist arthroplasty*

Occupational therapy for rheumatoid arthritis.
Cochrane Database of Systematic Reviews, no. 1 (2004)
http://onlinelibrary.wiley.com/doi/10.1002/14651858.CD003114.pub2/abstract

Splints and orthosis for treating rheumatoid arthritis.
Cochrane Database of Systematic Reviews, no. 4 (2001)
http://onlinelibrary.wiley.com/doi/10.1002/14651858.CD004018/abstract

Post-operative therapy for metacarpophalangeal arthroplasty.
Cochrane Database of Systematic Reviews, no. 1 (2008)
http://onlinelibrary.wiley.com/doi/10.1002/14651858.CD003522.pub2/abstract

**Gout**

Interventions for tophi in gout.
Cochrane Database of Systematic Reviews, no. 10 (2014)
http://onlinelibrary.wiley.com/doi/10.1002/14651858.CD010069.pub2/abstract

**Distal radius fractures**

Conservative interventions for treating distal radial fractures in adults.
Cochrane Database of Systematic Reviews, no. 2 (2003)
http://onlinelibrary.wiley.com/doi/10.1002/14651858.CD000314/abstract

External fixation versus conservative treatment for distal radial fractures in adults.
Cochrane Database of Systematic Reviews, no. 3 (2007)
http://onlinelibrary.wiley.com/doi/10.1002/14651858.CD006194.pub2/abstract

Different methods of external fixation for treating distal radial fractures in adults.
Cochrane Database of Systematic Reviews, no. 1 (2008)
http://onlinelibrary.wiley.com/doi/10.1002/14651858.CD006522.pub2/abstract

Percutaneous pinning for treating distal radial fractures in adults.
Cochrane Database of Systematic Reviews, no. 3 (2007)
http://onlinelibrary.wiley.com/doi/10.1002/14651858.CD006080.pub2/abstract

Pin site care for preventing infections associated with external bone fixators and pins.
Cochrane Database of Systematic Reviews, no. 12 (2013)
http://onlinelibrary.wiley.com/doi/10.1002/14651858.CD004551.pub3/abstract

Closed reduction methods for treating distal radial fractures in adults.
Cochrane Database of Systematic Reviews, no. 1 (2003)
http://onlinelibrary.wiley.com/doi/10.1002/14651858.CD003763/abstract

Bone grafts and bone substitutes for treating distal radial fractures in adults.
Cochrane Database of Systematic Reviews, no. 2 (2008)
http://onlinelibrary.wiley.com/doi/10.1002/14651858.CD006836.pub2/abstract

Rehabilitation for distal radial fractures in adults.
Cochrane Database of Systematic Reviews, no. 9 (2015)
http://onlinelibrary.wiley.com/doi/10.1002/14651858.CD003324.pub3/abstract

Anaesthesia for treating distal radial fracture in adults.
Cochrane Database of Systematic Reviews, no. 3 (2002)
http://onlinelibrary.wiley.com/doi/10.1002/14651858.CD003320/abstract

**Carpal pathology**

Computed tomography versus magnetic resonance imaging versus bone scintigraphy for clinically suspected scaphoid fractures in patients with negative plain radiographs.
Cochrane Database of Systematic Reviews, no. 6 (2015)
http://onlinelibrary.wiley.com/doi/10.1002/14651858.CD010023.pub2/abstract

**Metacarpal fractures & injuries**

Conservative treatment for closed fifth (small finger) metacarpal neck fractures.
Cochrane Database of Systematic Reviews, no. 3 (2005)
http://onlinelibrary.wiley.com/doi/10.1002/14651858.CD003210.pub3/abstract

**Digital fractures & injuries**

Conservative interventions for treating hyperextension injuries of the proximal interphalangeal joints of the fingers.
Cochrane Database of Systematic Reviews, no. 2 (2013)
http://onlinelibrary.wiley.com/doi/10.1002/14651858.CD009030.pub2/abstract

Interventions for treating mallet finger injuries.
Cochrane Database of Systematic Reviews, no. 3 (2004)
http://onlinelibrary.wiley.com/doi/10.1002/14651858.CD004574.pub2/abstract

Antibiotics for preventing infection in open limb fractures.
Cochrane Database of Systematic Reviews, no. 1 (2004)
http://onlinelibrary.wiley.com/doi/10.1002/14651858.CD003764.pub2/abstract
Full text has phalangeal fractures in the hand as a planned subgroup for the analysis, and three included studies were limited to open fractures of fingers.

**Microvascular reconstruction**

Low molecular weight heparin for prevention of microvascular occlusion in digital replantation.
Cochrane Database of Systematic Reviews, no. 7 (2013)
http://onlinelibrary.wiley.com/doi/10.1002/14651858.CD009894.pub2/abstract

**Complex regional pain syndrome and neuropathic pain**

Interventions for treating pain and disability in adults with complex regional pain syndrome- an overview of systematic reviews.
Cochrane Database of Systematic Reviews, no. 4 (2013)
http://onlinelibrary.wiley.com/doi/10.1002/14651858.CD009416.pub2/abstract

Local anaesthetic sympathetic blockade for complex regional pain syndrome.
Cochrane Database of Systematic Reviews, no. 7 (2016)
http://onlinelibrary.wiley.com/doi/10.1002/14651858.CD004598.pub4/abstract

Physiotherapy for pain and disability in adults with complex regional pain syndrome (CRPS) types I and II.
Cochrane Database of Systematic Reviews, no. 2 (2016)
http://onlinelibrary.wiley.com/doi/10.1002/14651858.CD010853.pub2/abstract

**Other hand trauma**

Vocational rehabilitation for enhancing return-to-work in workers with traumatic upper limb injuries.
Cochrane Database of Systematic Reviews, no. 10 (2013)

http://onlinelibrary.wiley.com/doi/10.1002/14651858.CD010002.pub2/abstract

Antibiotic prophylaxis for mammalian bites.
Cochrane Database of Systematic Reviews, no. 2 (2001)
http://onlinelibrary.wiley.com/doi/10.1002/14651858.CD001738/abstract
Full text indicates hand bites included, and included studies and conclusions include hand-specific content and differences from other bite sites.

Primary closure versus delayed closure for non bite traumatic wounds within 24 hours post injury
Cochrane Database of Systematic Reviews, no. 10 (2013)
http://onlinelibrary.wiley.com/doi/10.1002/14651858.CD008574.pub3/abstract

**Anaesthesia and analgesia**

Adrenaline with lidocaine for digital nerve blocks.
Cochrane Database of Systematic Reviews, no. 3 (2015)
http://onlinelibrary.wiley.com/doi/10.1002/14651858.CD010645.pub2/abstract

**Protocols**

*n = 18*

**Infection & antibiotic use**

Peri-operative antibiotics for hand trauma involving tendons and nerves.
Cochrane Database of Systematic Reviews, no. 6 (2012)
http://onlinelibrary.wiley.com/doi/10.1002/14651858.CD002107.pub2/abstract

**Carpal tunnel syndrome**

Acupuncture and related interventions for the treatment of symptoms associated with carpal tunnel syndrome.
Cochrane Database of Systematic Reviews, no. 7 (2014)
http://onlinelibrary.wiley.com/doi/10.1002/14651858.CD011215/abstract

Open release for carpal tunnel syndrome.
Cochrane Database of Systematic Reviews, no. 3 (2014)
http://onlinelibrary.wiley.com/doi/10.1002/14651858.CD011041/abstract

Absorbable versus non-absorbable sutures for carpal tunnel release.
Cochrane Database of Systematic Reviews, no. 6 (2015)
http://onlinelibrary.wiley.com/doi/10.1002/14651858.CD011757/abstract

**Stroke & adult spasticity**

Assistive technology, including orthotic devices, for the management of contractures in adult stroke patients.
Cochrane Database of Systematic Reviews, no. 10 (2013)
http://onlinelibrary.wiley.com/doi/10.1002/14651858.CD010779/abstract

Physical treatment interventions for managing spasticity after stroke.
Cochrane Database of Systematic Reviews, no. 7 (2011)
http://onlinelibrary.wiley.com/doi/10.1002/14651858.CD009188/abstract

**Other nerve disorders**

*Dystonia*

Botulinum a toxin for focal hand dystonia.
Cochrane Database of Systematic Reviews, no. 4 (2015)

http://onlinelibrary.wiley.com/doi/10.1002/14651858.CD004556.pub2/abstract

**Dupuytren's contracture**

Rehabilitation after surgery for Dupuytren's contracture.
Cochrane Database of Systematic Reviews, no. 2 (2007)
http://onlinelibrary.wiley.com/doi/10.1002/14651858.CD006508/abstract

**Trigger digits**

Surgery for trigger finger.
Cochrane Database of Systematic Reviews, no. 6 (2012)
http://onlinelibrary.wiley.com/doi/10.1002/14651858.CD009860/abstract

**Osteoarthritis of the hand**

Exercises for hand osteoarthritis.
Cochrane Database of Systematic Reviews, no. 2 (2013)
http://onlinelibrary.wiley.com/doi/10.1002/14651858.CD010388/abstract

**Inflammatory conditions and arthropathies**

*Rheumatoid arthritis & wrist arthroplasty*

Exercise therapy for the rheumatoid hand.
Cochrane Database of Systematic Reviews, no. 4 (2012)
http://onlinelibrary.wiley.com/doi/10.1002/14651858.CD003832.pub2/abstract

**Space-occupying lesions**

*Ganglia*

Interventions for ganglion cysts in adults.
Cochrane Database of Systematic Reviews, no. 2 (2005)
http://onlinelibrary.wiley.com/doi/10.1002/14651858.CD005327/abstract

**Distal radius fractures**

Internal fixation for treating distal radius fractures in adults.
Cochrane Database of Systematic Reviews, no. 7 (2014)
http://onlinelibrary.wiley.com/doi/10.1002/14651858.CD011213/abstract

Internal fixation versus other surgical methods for treating distal radius fractures in adults.
Cochrane Database of Systematic Reviews, no. 7 (2014)
http://onlinelibrary.wiley.com/doi/10.1002/14651858.CD011212/abstract

**Carpal pathology**

*Scaphoid fractures*

Surgical versus conservative interventions for treating acute scaphoid fractures in adults.
Cochrane Database of Systematic Reviews, no. 6 (2014)
http://onlinelibrary.wiley.com/doi/10.1002/14651858.CD010035.pub2/abstract

Conservative interventions for treating scaphoid fractures in adults.
Cochrane Database of Systematic Reviews, no. 8 (2013)
http://onlinelibrary.wiley.com/doi/10.1002/14651858.CD010713/abstract

**Digital fractures & injuries**

Interventions for treating ulnar collateral ligament injuries of the thumb.
Cochrane Database of Systematic Reviews, no. 8 (2014)
http://onlinelibrary.wiley.com/doi/10.1002/14651858.CD011267/abstract

**Other hand trauma**

*Complex regional pain syndrome and neuropathic pain*

Vitamin C for preventing complex regional pain syndrome (Type I) after wrist fractures in adults.
Cochrane Database of Systematic Reviews, no. 12 (2015)
http://onlinelibrary.wiley.com/doi/10.1002/14651858.CD007817.pub2/abstract

**Guidelines**

*n= 16*

**Trigger digits**

BSSH: Evidence based management of adult trigger digits

**Stroke & adult spasticity**

NICE: Stroke rehabilitation in adults : guidance (CG162)

SIGN: Guideline 118: Management of patients with stroke: rehabilitation, prevention and management of complications, and discharge planning - Full guideline [PDF]

**Dupuytren's contracture**

NICE: Radiation therapy for early Dupuytren's disease - guidance (IPG368)

NICE: Needle fasciotomy for Dupuytren's contracture - guidance (IPG43)

**Osteoarthritis**

NICE: Osteoarthritis: care and management : guidance (CG177)

NICE: Artificial metacarpophalangeal and interphalangeal joint replacement for end-stage arthritis - guidance (IPG110)

NICE: Artificial trapeziometacarpal joint replacement for end-stage osteoarthritis - guidance (IPG111)

**Rheumatoid arthritis & wrist arthroplasty**

NICE: Rheumatoid arthritis in adults: management : guidance (CG79)

NICE: Total wrist replacement - guidance (IPG271)

**Hand fractures**

NICE: Fractures (non-complex): assessment and management : guidance (NG38)

NICE: Fractures (complex): assessment and management : guidance (NG37)

NICE: [Low-intensity pulsed ultrasound to promote fracture healing - guidance (IPG374)]

**Hand transplantation**

NICE: [Hand allotransplantation - guidance (IPG383)]

**Prostheses**

NICE: [Direct skeletal fixation of limb or digit prostheses using intraosseous transcutaneous implants - guidance (IPG270)]

**Brachial plexus injury**

NICE: [Phrenic nerve transfer in brachial plexus injury - guidance (IPG468)]
