## Supplemental file 3 for "Uncertainty extraction and verification for the Common Conditions Affecting the Hand and Wrist Priority Setting Partnership: A proposed methodology for Priority Setting Partnerships with the James Lind Alliance"

| Source<br>(Review/protocol or guideline reference) | Reason for evidence gap (see Key) | Population (P) | Intervention (I) | Comparator ( C ) | Outcomes (O) as per Key | Free text ofuncertainty(ies) in PICO format whenever possible | Original verbatim Uncertainty<br>(extracted from review/ protocol or guideline text) | Corresponding Stakeholder Question |
|---|---|---|---|---|---|---|---|---|
| Open release for carpal tunnel syndrome.<br>Cochrane Database of Systematic Reviews, no. 3 (2014)<br>http://onlinelibrary.wiley.com/doi/10.1002/14651858.CD011041/abstract | B | participants who have a clinical diagnosis of CTS with or without electrophysiological confirmation. We will accept the authors' definition of CTS and their views of what constituted electrophysiological confirmation. | Open CTD | endoscopic carpal tunnel release (ECTR), release with mini-open techniques (MOCTR) or OCTR techniques with concomitant interventions (such as lengthening of flexor retinaculum, internal neurolysis, epineurotomy or tenosynovectomy). | Symptoms, function, complications, time off work | In people with a clinical history of carpalt tunnel syndrome is any surgical intervention superior to standard open carpal tunnel release | Protocol so cannot conclude | |
| Absorbable versus non-absorbable sutures for carpal tunnel release.<br>Cochrane Database of Systematic Reviews, no. 6 (2015)<br>http://onlinelibrary.wiley.com/doi/10.1002/14651858.CD011757/abstract | B | Adaults undergoing carpal tunnel release surgery. | Nonabsorbable suture | Absorbable suture | Pain, function scar and complications | In adults undergoing carpal tunnel surgery is wound closure with non-absorbable or absorbable sutures superior | Protocol so cannot conclude | Does the surgical method (i.e. the exact technique used) influence the outcome following surgery for peripheral nerve compression (for example carpal tunnel syndrome or cubital tunnel syndrome)? |
| Surgery for trigger finger.<br>Cochrane Database of Systematic Reviews, no. 6 (2012)<br>http://onlinelibrary.wiley.com/doi/10.1002/14651858.CD009860/abstract | B | Adults with trigger finger listed for surgery | Surgical release (open, percutaneous, endoscopic) | Placebo, non-operative, other | Resolution of trigering, tenderness, function | Unclear - could be is surgery for tribgger finger superior to other treatments or could be which type of surgery is best for trigger finger? | Protocol so cannot conclude | In the treatment of non-traumatic tendon problems in the hand and wrist, how does surgical intervention compare to non-surgical methods? |
| Surgical versus conservative interventions for treating acute scaphoid fractures in adults.<br>Cochrane Database of Systematic Reviews, no. 6 (2014)<br>http://onlinelibrary.wiley.com/doi/10.1002/14651858.CD010035.pub2/abstract | B | Protocol withdrawn | | | | | | N/A |
| Conservative interventions for treating scaphoid fractures in adults.<br>Cochrane Database of Systematic Reviews, no. 8 (2013)<br>http://onlinelibrary.wiley.com/doi/10.1002/14651858.CD010713/abstract | B | Adults with scaphoid fractures | One type of plaster cast | Another type of plaseter cast (thumb incorporated, wrist position to vary) | Function, complications, fracture healing, quality of life | Which type of plaster cast is best for treating adults with scaphoid fractures | Protocol so cannot conclude | Which patients with a recent scaphoid fracture would benefit from surgery rather than cast or splint treatment? |
| | | | Conservative | Conservative | Resolution of triggering at 1 month | | | |
| BSSH: Evidence based management of adult trigger digits | A | Adults with trigger finger without Rheumatoid Arthritis | Operative | Other operative | Resolution of triggering | • surgery versus corticosteroid injections with outcomes measured beyond four months;<br>• corticosteroid versus corticosteroid combined with local anaesthetic<br>• DIP joint and MCP joint splints with corticosteroid injection;<br>• individual hand therapy treatment modalities.<br>• Treatment of trigger fingers in those with rheumatoid arthritis and diabetes<br>mellitus<br>• Treatment strategies involving more than one injection containing steroid (i.e.<br>giving a second or even third steroid injection) | Which patients should be referred to hand surgeons?<br>• Which treatments are superior to other treatments?<br>• Which treatments are more cost-effective than other treatments?<br>• What treatments should be offered to patients?<br>• At what clinical stage should different treatments be offered to patients?<br>• What outcomes can be expected from particular treatments?<br>• What future research might be beneficial in clarifying optimal treatment? | In the treatment of non-traumatic tendon problems in the hand and wrist, how does surgical intervention compare to non-surgical methods? |
| | | | Operative | Conservative | Resolution of triggering | | | In the treatment of non-traumatic tendon problems in the hand and wrist, how does surgical intervention compare to non-surgical methods? |
| NICE: Artificial trapeziometacarpal joint replacement for end-stage osteoarthritis - guidance (IPG111) | A | Adults with end-stage osteoarthritis undergoing surgery. | Implant operative | Other operative | NO RCTs | Concluded implant arthroplasties are a safe treatment option - no key uncertainty | Whether implant arthroplasties are a safe procedure | Regarding patient and cost benefits, are movement preserving surgeries, such as joint replacement or cartilage replacement, preferential to fusion (permanent stiffening) in the treatment of painful joints in the hand/wrist? |
| Surgery for Dupuytren's contracture of the fingers.<br>Cochrane Database of Systematic Reviews, no. 12 (2015)<br>http://onlinelibrary.wiley.com/doi/10.1002/14651858.CD010143.pub2/abstract | E | Adults with Dupuytren's contractures of the finger(s) - not thumb | Any surgery | Control, non-operative management, other surgery | Function, clinical, complications | Currently, insufficient evidence is available to show the relative superiority of different surgical procedures (needle fasciotomy vs fasciectomy, or interposition firebreak skin grafting vs z-plasty closure of fasciectomy). Low-quality evidence suggests that postoperative splinting may not improve outcomes and may impair outcomes by reducing active flexion. | Comparative analysis of the outcomes of different surgical treatment options for Dupuytren's disease is needed to investigate whether more invasive procedures, such as dermofasciectomy, have lower 'recurrent contracture' rates, and whether any such benefit is outweighed by a higher rate of adverse events (complications) or an unacceptably longer or more difficult rehabilitation period. | In patients with Dupuytren's disease, what invasive techniques give the best results in terms of function and recurrence? |
| Surgical treatment options for carpal tunnel syndrome.<br>Cochrane Database of Systematic Reviews, no. 4 (2007)<br>http://onlinelibrary.wiley.com/doi/10.1002/14651858.CD003905.pub3/abstract | E | people with a clinical diagnosis of CTS with or without electrophysiological confirmation | any of: standard open carpal tunnel release (OCTR); endoscopic carpal tunnel release (ECTR);<br>open carpal tunnel release with additional procedures; and open carpal tunnel release using various incision techniques | any of: standard open carpal tunnel release (OCTR); endoscopic carpal tunnel release (ECTR);<br>open carpal tunnel release with additional procedures; and open carpal tunnel release using various incision techniques | Function, clinical, complications | There is no strong evidence supporting the need for replacement of standard open carpal tunnel release by existing alternative surgical procedures for the treatment of carpal tunnel syndrome. | Various surgical techniques are available and there has been considerable discussion about which method is the most effective. | |
| Surgical versus non-surgical treatment for carpal tunnel syndrome.<br>Cochrane Database of Systematic Reviews, no. 4 (2008)<br>http://onlinelibrary.wiley.com/doi/10.1002/14651858.CD001552.pub2/abstract | E | Patients diagnosed with CTS | All surgical techniques | All non-surgical treatments | Clinical, Functional and complications | Surgical treatment of carpal tunnel syndrome relieves symptoms significantly better than splinting. Further research is needed to discover whether this conclusion applies to people with mild symptoms and whether surgical treatment is better than steroid injection. | There is no universally accepted therapy for CTS (Rosenbaum 1993) although clinical guidelines have been suggested (AAN 1993). For symptomatic patients a range of treatment is offered varying widely around the world, within individual countries, and even hospitals. Most patients are treated non surgically (Miller 1994). | When is surgery of a greater benefit than non-surgical methods in the treatment of peripheral nerve compression (for example carpal or cubital tunnel syndrome)? |
| Endoscopic release for carpal tunnel syndrome.<br>Cochrane Database of Systematic Reviews, no. 1 (2014)<br>http://onlinelibrary.wiley.com/doi/10.1002/14651858.CD008265.pub2/abstract | E | Patients with clinical diagnosis of CTS with or without electrophysiological confirmation. | Endoscopic CTR | Other surgical treatments (Open release and its variations) | Clinical, Functional and complications | In this review, with support from low quality evidence only, OCTR and ECTR for carpal tunnel release are about as effective as each other in relieving symptoms and improving functional status, although there may be a functionally significant benefit of ECTR over OCTR in improvement in grip strength. ECTR appears to be associated with fewer minor complications compared to OCTR, but we found no difference in the rates of major complications. Return | The main questions that need to be answered relate to its efficacy and safety compared to OCTR, which remains the gold standard method for carpal tunnel release. | |
| Splinting for carpal tunnel syndrome.<br>Cochrane Database of Systematic Reviews, no. 7 (2012)<br>http://onlinelibrary.wiley.com/doi/10.1002/14651858.CD010003/abstract | D | All participants with a diagnosis of CTS, as defined by the authors of each study. Excluded if past history of Carpal tunnel surgery | All splinting interventions. | No treatment, placebo and other non-surgical interventions. | Clinical, Functional and complications | Overall, there is limited evidence that a splint worn at night is more effective than no treatment in the short term, but there is insufficient evidence regarding the effectiveness and safety of one splint design or wearing regimen over others, and of splint over other non-surgical interventions for CTS. More research is needed on the long-term effects of this intervention for CTS. | Splinting is usually offered to people with mild to moderate symptoms. However, the effectiveness and duration of the benefit of splinting for this condition remain unknown. | What are the most effective non-surgical methods of treating peripheral nerve compression (for example carpal tunnel syndrome or cubital tunnel syndrome)? |
| NICE: Total wrist replacement - guidance (IPG271) | A | Patients with arthritic wrists | Total wrist replacement | No treatment, wrist arthrodesis, | Safety and efficacy | Does total wrist replacement offer a safe and effective option for the treatment of arthritic wrists? | No long term studies | Regarding patient and cost benefits, are movement preserving surgeries, such as joint replacement or cartilage replacement, preferential to fusion (permanent stiffening) in the treatment of painful joints in the hand/wrist? AND When should patients with pain or deformity following joint damage or arthritis undergo surgery to correct/improve it? |
| NICE: Artificial metacarpophalangeal and interphalangeal joint replacement for end-stage arthritis - guidance (IPG110) | A | Patients with osteoarthritis of hand affecting MCP and/or IP joints | Artificial joint replacment | Conservative treatments, excision, interposition, arthrodesis | Safety and efficacy | Does artificial joint replacement in the fingers offer a safer or more effective option than other surgical and non-surgical methods? | No good studies; limited search terms in available systematic reviews which may bias results | Regarding patient and cost benefits, are movement preserving surgeries, such as joint replacement or cartilage replacement, preferential to fusion (permanent stiffening) in the treatment of painful joints in the hand/wrist? AND When should patients with pain or deformity following joint damage or arthritis undergo surgery to correct/improve it? |
| Conservative interventions for treating distal radial fractures in adults.<br>Cochrane Database of Systematic Reviews, no. 2 (2003)<br>http://onlinelibrary.wiley.com/doi/10.1002/14651858.CD000314/abstract | D | Skeletally mature patients with distal radius fracture | Conservative treatment methods | Other treatments or placebo | Anatomical, functional, clinical outcomes and resource use | What is the best non-surgical treatment for distal radial fractures? | Given that a distal radius fracture in adults is a common injury and given that there is very limited knowledge about the best method of treatment, either conservative or surgical (Handoll 2005a), further research is called for. | Can alternatives to plaster casts be used in the treatment of wrist fractures? AND What patient and injury factors predict outcome following wrist fracture and determine which patients may benefit from the various treatment options? |
| Corticosteroid injection for trigger finger in adults.<br>Cochrane Database of Systematic Reviews, no. 1 (2009)<br>http://onlinelibrary.wiley.com/doi/10.1002/14651858.CD005617.pub2/abstract | E | Adults (>18) with clinical diagnosis of trigger finger (non-infectious origin) | Injectable corticosteroids | Placebo or non-steroid injection, NSAIDs/medications, splints, surgery, combination treatments or no intervention | Success (complete, number of trigegring episodes), functional status, side effects, pain | Is corticosteroid injection better in the treatment of trigger finger than other non-surgical, medical or surgical interventions including placebos? | | In the treatment of non-traumatic tendon problems in the hand and wrist, how does surgical intervention compare to non-surgical methods? |
| Computed tomography versus magnetic resonance imaging versus bone scintigraphy for clinically suspected scaphoid fractures in patients with negative plain radiographs.<br>Cochrane Database of Systematic Reviews, no. 6 (2015)<br>http://onlinelibrary.wiley.com/doi/10.1002/14651858.CD010023.pub2/abstract | E | Patients with a possible scaphoid fracture | CT, MRI | CT, MRI | Correct diagnosis, economic impact, PROM | What is the best method of detecting the occult or suspected scaphoid fracture? | Lack of comparitive studies. Prospective studies, perhaps involving randomisation of diagnostic tests, with direct comparisons of CT and MRI in the same patient population would add valuable data. We question the need for further research evaluating BS because of its limited use and invasive character. It would be useful if such studies incorporated economic (direct and indirect costs) and patient-related outcome measures (e.g. Disabilities of the Arm Shoulder and Hand, Patient Related Wrist Evaluation). Given the debate on the current best available reference standard (six week radiographs), consideration should be given to the practicalities of a check radiological follow-up, perhaps at one year, to examine for missed fractures. Prior to these, studies looking at ways to improve initial diagnostic management are needed | |
| Local corticosteroid injection for carpal tunnel syndrome.<br>Cochrane Database of Systematic Reviews, no. 2 (2007) | D | Uni or bilateral CTS with no history of CT release diagnosed in line with AAN 1993 standards | Corticosteroid injection in or near the carpal tunnel | Placebo injection, systemic steroid, splints/NSAIDs, high dose steroid injection, laser treatment, short acting steroid, repreated injection, location of injection, ionto-phonophoresis | Clinical improvement at 3/12; clinical improvement at before 3/12 or at 1 year, neurphysiological improvement, requirement for surgery, QOL, return to work | Does corticosteroid injection have long-term efficacy and cost effectivemness in the treatment of CTS compared to other non-surgical treatments? | Research is required to determine the duration of benefit from local corticosteroid injection and to identify candidates for treatment based on severity and duration of symptoms. Local corticosteroid injection should also be compared to, and combined with, other non-surgical and even surgical interventions to determine the optimum management of CTS. | When is surgery of a greater benefit than non-surgical methods in the treatment of peripheral nerve compression (for example carpal or cubital tunnel syndrome)? |

| | | | | | | | | |
|---|---|---|---|---|---|---|---|---|
| http://onlinelibrary.wiley.com/doi/10.1002/14651858.CD001554.pub2/abstract | E (for injection v placebo; low v high) otherwise D | Uni or bilateral CTS with no history of CT release diagnosed in line with AAN 1993 standards | Corticosteroid injection in or near the carpal tunnel | Placebo injection, systemic steroid, splints/NSAIDs, high dose steroid injection, laser treatment, short acting steroid, repreated injection, location of injection, ionto-phonophoresis | Clinical improvement at 3/12; clinical improvement at before 3/12 or at 1 year, neurphysiological improvement, requirement for surgery, QOL, return to work | What is the best method of steroid delivery in the treatment of CTS? | Research is required to determine the duration of benefit from local corticosteroid injection and to identify candidates for treatment based on severity and duration of symptoms. Local corticosteroid injection should also be compared to, and combined with, other non-surgical and even surgical interventions to determine the optimum management of CTS. | When is surgery of a greater benefit than non-surgical methods in the treatment of peripheral nerve compression (for example carpal or cubital tunnel syndrome)? |
| Non-surgical treatment (other than steroid injection) for carpal tunnel syndrome.<br>Cochrane Database of Systematic Reviews, no. 1 (2003)<br>http://onlinelibrary.wiley.com/doi/10.1002/14651858.CD003219/abstract | E (for oral steroid v placebo; USS v placebo) otherwise D | Clinical diagnosis of CTS | Non-surgical methods (night splint, full time v night, neutral v extended,USS, ergonomics, diuretics, NSAIDs, orla steroid, Vit B6, nerve/tendon glides, yoga v splint, neurodynamic mobilisation, carpal bone manipulation, chirpractic, lser, steroid/insulin injection | Other non-surgical methods or placebo | QOL, objective measures, neurophysiological measures, clinical improvement 3/12 and 12/12, need for surgery | Which is the best method of non-surgical treatment in the long-term resolution of CTS? | More high quality research is needed to strengthen the moderate to limited evidence currently available on non-surgical treatment. Future research needs to examine the relative contributions of different non-surgical treatments for CTS, the optimal forms of delivery, the duration of any benefit (both during active treatment and after treatment cessation) and the optimal timing of delivery during the course of CTS. More high quality studies are required to establish better evidence to direct clinical practice. | When is surgery of a greater benefit than non-surgical methods in the treatment of peripheral nerve compression (for example carpal or cubital tunnel syndrome)? |
| Therapeutic ultrasound for carpal tunnel syndrome.<br>Cochrane Database of Systematic Reviews, no. 3 (2013)<br>http://onlinelibrary.wiley.com/doi/10.1002/14651858.CD009601.pub2/abstract | D | Clinical diagnosis of CTS without previous surgery | therapeutic ultrasound | | Any short term improvement (subjective, outcome, satisfaction), adverse effects, improvement (pain, apraesthesia, night syx) at short <3/12 or lolng >3/12; QOL | Is therapeutic ultrasound safe and effective in the treatment of CTS? | Large scale, methodologically rigorous randomised trials are needed to assess the safety and efficacy of different therapeutic ultrasound regimens as compared with other non-surgical interventions for CTS. | When is surgery of a greater benefit than non-surgical methods in the treatment of peripheral nerve compression (for example carpal or cubital tunnel syndrome)? |
| | E | People with clinical symptoms suggesting the presence of UNE with or without neurophysiological evidence of entrapment. | Surgery (Simple decompression) | Surgery (all types of transposition) | Functional, clinical, complications, quality of life | | | When is surgery of a greater benefit than non-surgical methods in the treatment of peripheral nerve compression (for example carpal or cubital tunnel syndrome)? |
| Treatment for ulnar neuropathy at the elbow.<br>Cochrane Database of Systematic Reviews, no. 11 (2016)<br>http://onlinelibrary.wiley.com/doi/10.1002/14651858.CD006839.pub3/abstract | C | People with clinical symptoms suggesting the presence of UNE with or without neurophysiological evidence of entrapment. | Surgery | Conservative treatments | Functional, clinical, complications, quality of life | In people with clinical symptoms of ulnar neuropathy at the elbow, what is the best surgical or conservative treatment .....in terms of functional, clinical outcomes, complications, quality of life? | "The available evidence is not sufficient to identify the best treatment based on clinical, neurophysiological, and imaging characteristics." "No evidence is available on the effects of surgery on quality of life and imaging characteristics of the ulnar nerve at the elbow."<br>"Future research in this area should include RCTs to evaluate the effectiveness of conservative treatments." "Moreover, there is a need for a trial comparing conservative and surgical treatment." | When is surgery of a greater benefit than non-surgical methods in the treatment of peripheral nerve compression (for example carpal or cubital tunnel syndrome)? |
| | D | People with clinical symptoms suggesting the presence of UNE with or without neurophysiological evidence of entrapment. | Conservative treatment | Conservative treatment | Functional, clinical, complications, quality of life | | | When is surgery of a greater benefit than non-surgical methods in the treatment of peripheral nerve compression (for example carpal or cubital tunnel syndrome)? |
| Radiation therapy for early Dupuytren's disease: Interventional procedures guidance [IPG573] | A | Adults with Dupuytrens Disease | radiation for Dupuytrens | | Functional, Clinical, complications Quality of life.Absence of progression, time to recurrence or progression to a functionally significant contracture, and rates of subsequent surgery | NICE enourage further research inc RCT | Current evidence on its efficacy is inadequate in quantity and quality, and is difficult to interpret because of uncertainty about the natural history of Dupuytren's disease. Procedure should only be used with special arrangements for clinical governance, consent and audit or research | What non-surgical treatments have benefits over surgery in the treatment of Dupuytren's disease? |
| NICE: Needle fasciotomy for Dupuytren's contracture - guidance (IPG43) | A | Adults with Dupuytrens of one or more digits | needle fasciotomy | collagenase, surgery, nothing | Functional, Clinical. finger flexion, recurrence | what is the long term efficacy and cost | | |
| NICE: Phrenic nerve transfer in brachial plexus injury - guidance (IPG468) | A | Adult brachial plexus injury patients with transection of nerve. Not recommended in children | Phrenic nerve transfer | direct repair, muscle transfer, nerve transfer, tendon transfer, nerve graft | Functional, Clinical, complications Quality of life. Arm function, long-term functional and quality-of-life outcomes, and evidence on safety. | No evidence of long term functional advantage or safety compared to other techniques | | What interventions/treatments will have the most positive effect following nerve injury? |
| NICE: Hand allotransplantation - guidance (IPG383) | A | patients who have undergone upper limb amputation | Hand Transplant | prosthetics, doing nothing | Functional, Clinical, complications Quality of life. graft/host survival. Cancer risk. Function. Pain. Return to work | Is this procedure safe and does it offer a functional improvement over bionics | Current evidence on the efficacy and safety of hand allotransplantation is inadequate in quantity | Does prosthetic (artificial) replacement or hand transplantation have the greater clinical and cost benefits when used in the hand and wrist? |
| Treatment for thoracic outlet syndrome.<br>Cochrane Database of Systematic Reviews, no. 11 (2014)<br>http://onlinelibrary.wiley.com/doi/10.1002/14651858.CD007218.pub3/abstract | D | patients with thoracic outlet syndrome | opertaive | non-operative treatment | Functional, Clinical, complications Quality of life.The primary outcome pain rating at least six months after the intervention.Secondary outcomes change in muscle strength, disability, experiences of paresthesias and adverse effects interventions | need to define diagnosis and outcome measure. RCT of treatment vs no treatement and between each other | Need for agreed definition for the diagnosis of TOS, especially the disputed form, agreed outcome measures, and high quality randomized trials that compare the outcome of interventions with no treatment and with each other. | When is surgery of a greater benefit than non-surgical methods in the treatment of peripheral nerve compression (for example carpal or cubital tunnel syndrome)? |
| Low molecular weight heparin for prevention of microvascular occlusion in digital replantation.<br>Cochrane Database of Systematic Reviews, no. 7 (2013)<br>http://onlinelibrary.wiley.com/doi/10.1002/14651858.CD009894.pub2/abstract | D | patients undergoing digital replant | Low molecular weight heaptin | Unfractionated heparin | clinical, complications | whether LMWH or UFH is better. No trial comparing heparin to no-heparin | Further well-designed and adequately powered clinical trials are warranted. | |
| Surgical treatment for the thumb-in-palm deformity in patients with cerebral palsy.<br>Cochrane Database of Systematic Reviews, no. 4 (2005)<br>http://onlinelibrary.wiley.com/doi/10.1002/14651858.CD004093.pub2/abstract | C | thumb-in-palm deformity in patients with spastic cerebral palsy | efficacy of surgery, selection criteria for surgery, Outcomes | | Functional, Clinical, complications Quality of life | what is the benefit of surgery? What are the selection criteria? | This review has demonstrated the need for randomized clinical trials or controlled clinical trials on the surgical treatment of thumb-in-palm deformity. | Which patients with paralysis, spasticity or functional loss in the upper limb following brain or nerve injury, benefit from surgery in addition to non-surgical treatments? |
| Corticosteroid injection for de Quervain's tenosynovitis.<br>Cochrane Database of Systematic Reviews, no. 3 (2009)<br>http://onlinelibrary.wiley.com/doi/10.1002/14651858.CD005616.pub2/abstract | C | Pregnant lactating women with De Quervains | steroid | splint | clinical, complications | Are steroids effective in reducing pain in patinets with De Quervains caompared to placebo? | Uncertain whether Corticosteroid injections reduces pain because of the very low quality of the evidence | In the treatment of non-traumatic tendon problems in the hand and wrist, how does surgical intervention compare to non-surgical methods? |
| Antibiotic prophylaxis for mammalian bites.<br>Cochrane Database of Systematic Reviews, no. 2 (2001)<br>http://onlinelibrary.wiley.com/doi/10.1002/14651858.CD001738/abstract | E | Patientes with bites from mammals | Antibiotic | placeabo / non intervention | clinical, complications | RCT to determine evidence for use of antibiotics follwing dog bites. Better RCT to examine antibiotic use in hand bites | There is evidence from one trial that prophylactic antibiotics reduces the risk of infection after human bites but confirmatory research is required. There is no evidence that the use of prophylactic antibiotics is effective for cat or dog bites. There is evidence that the use of antibiotic prophylactic after bites of the hand reduces infection but confirmatory research is required. | Does the use of antibiotics affect the infection rate following hand trauma or elective hand surgery? |
| Primary closure versus delayed closure for non bite traumatic wounds within 24 hours post injury<br>Cochrane Database of Systematic Reviews, no. 10 (2013)<br>http://onlinelibrary.wiley.com/doi/10.1002/14651858.CD008574.pub3/abstract | C | Patients with non-bite traumatic wounds presenting within 24 hours | Primary closure | delayed closure | Clinical, complications Quality of life | RCT of non bite traumatic wound of primary versus dleayed closure. Outcome measures need to be defined. | There is currently no systematic evidence to guide clinical decision-making regarding the timing for closure of traumatic wounds. There is a need for robust research to investigate the effect of primary closure compared with delayed closure for non bite traumatic wounds presenting within 24 hours of injury | |
| Rehabilitation after surgery for Dupuytren's contracture.<br>Cochrane Database of Systematic Reviews, no. 2 (2007)<br>http://onlinelibrary.wiley.com/doi/10.1002/14651858.CD006508/abstract | B | Patients who havve had surgery for Duuytrens | Splinting | no splinting or alternate splinting regimes | Maintainence of surgical corrction, function, pain, range of movement, grip strength, quality of life | RCT of splint 1 v splint 2 (or no splint). This is an old protocol, and there have been RCTs in this area so may not be relevant | What post operative rehabilitation interventions are effective and safe for maintaining corrections achieved following surgical release of Dupuytren's contracture?; What post-operative rehabilitation interventions are effective for improving and maintaining function, reducing pain, achieving maximum range of movement and reducing adverse effects; What post-operative rehabilitation interventions are effective for improving grip and pinch grip strength and improving quality of life. | In previously treated Dupuytren's disease, do any additional techniques help prevent recurrence? |
| Botulinum a toxin for focal hand dystonia.<br>Cochrane Database of Systematic Reviews, no. 4 (2015)<br>http://onlinelibrary.wiley.com/doi/10.1002/14651858.CD004556.pub2/abstract | B - no abstract or protocol has been written | | | | | | | I would say this is rare and therefore not in scope. |
| Interventions for ganglion cysts in adults.<br>Cochrane Database of Systematic Reviews, no. 2 (2005)<br>http://onlinelibrary.wiley.com/doi/10.1002/14651858.CD005327/abstract | B | Adults with ganglion cysts | surgery | other treatmetn options (aspiration, steroid injection, lidocaine infiltration, hypertonic saline infiltration, hyaluronidase infiltration, traumatic destruction) | Quality of life, complications, recurrence | RCT of surgery v other intervention | To determine the efficacy and safety of different treatments (surgery, multiple punctures, aspiration, steroid infiltration, lidocaine infiltration, hypertonic saline infiltration, hyaluronidase infiltration and traumatic destruction) of ganglion cysts in adults. | In the treatment of common hand conditions, such as peripheral nerve compression syndromes (for example carpal tunnel syndrome), ganglia or arthritis of the fingers/thumb/wrist, do surgical interventions have a demonstrable benefit in patient reported outcome when compared with non-surgical methods or placebo (sham) surgery? |
| Vitamin C for preventing complex regional pain syndrome (Type I) after wrist fractures in adults.<br>Cochrane Database of Systematic Reviews, no. 12 (2015)<br>http://onlinelibrary.wiley.com/doi/10.1002/14651858.CD007817.pub2/abstract | B - protocol withdrawn | | | | | | | N/A |
| Adrenaline with lidocaine for digital nerve blocks.<br>Cochrane Database of Systematic Reviews, no. 3 (2015)<br>http://onlinelibrary.wiley.com/doi/10.1002/14651858.CD010645.pub2/abstract | D | Patients having surgery on fingers | Lidocaine with adrenaline | Ldocaine without adrenaline | Duration of anaesthesia, adverse events, cost, post-operative pain relief | RCT of plain v with adrenaline | From the limited data available, evidence is insufficient to recommend use or avoidance of adrenaline in digital nerve blocks. The evidence provided in this review indicates that addition of adrenaline to lidocaine may prolong the duration of anaesthesia and reduce the risk of bleeding during surgery, although the quality of the evidence is low. We have identified the need for researchers to conduct large trials that focus on other important outcomes such as adverse events, cost analysis and duration of postoperative pain relief. | |

| | | | | | | | | |
|---|---|---|---|---|---|---|---|---|
| Interventions for tophi in gout.<br>Cochrane Database of Systematic Reviews, no. 10 (2014)<br>http://onlinelibrary.wiley.com/doi/10.1002/14651858.CD010069.pub2/abstract | D | Patients with gouty tophi of the fingers | Surgery | Pegloticase infusion | Quality of life, complications, recurrence | RCT of surgery v medical therapy | This study showed pegloticase is probably beneficial in the management of tophi in gout, in terms of resolution of tophi, but with a high risk of adverse infusion reactions. However, there is a need for more RCT data considering other interventions, including surgical removal of tophi. | |
| | D | Patients with thumb base arthritis | Surgery | Sham surgery or non-surgical treatment (analgesics/splint, exercises) | Quality of life, pain, complications | RCT of surgery v non-surgical treatment | We did not find any studies that compared surgery with sham surgery or surgery with non-surgical interventions. None of the included trials reported global assessment, quality of life, and revision or re-operation rates. | In the treatment of common hand conditions, such as peripheral nerve compression syndromes (for example carpal tunnel syndrome), ganglia or arthritis of the fingers/thumb/wrist, do surgical interventions have a demonstrable benefit in patient reported outcome when compared with non-surgical methods or placebo (sham) surgery? |
| Surgery for thumb (trapeziometacarpal joint) osteoarthritis.<br>Cochrane Database of Systematic Reviews, no. 2 (2015)<br>http://onlinelibrary.wiley.com/doi/10.1002/14651858.CD004631.pub4/abstract | | | | | | | | |
| | D | Patients with thumb base arthritis | First surgical technique | Second surgical technique | Quality of life, pain, complications | RCT of surgery 1 v surgery 2 | Furthermore, the included studies were not of high enough quality to provide conclusive evidence that the compared techniques provided equivalent outcomes. | In the treatment of common hand conditions, such as peripheral nerve compression syndromes (for example carpal tunnel syndrome), ganglia or arthritis of the fingers/thumb/wrist, do surgical interventions have a demonstrable benefit in patient reported outcome when compared with non-surgical methods or placebo (sham) surgery? |
| External fixation versus conservative treatment for distal radial fractures in adults.<br>Cochrane Database of Systematic Reviews, no. 3 (2007)<br>http://onlinelibrary.wiley.com/doi/10.1002/14651858.CD006194.pub2/abstract | D | Adults with dorsally displaced distal radius fractures | External fixation | plaster cast immobilisation | Functional outcome, re-intervention, Quality of life, pain, complications, cost | RCT of surgery v cast | There was insufficient evidence to confirm a superior overall functional or clinical result for the external fixation group. External fixation was associated with a high number of complications, such as pin-track infection, but many of these were minor. There was insufficient evidence to establish a difference between the two groups in serious complications such as reflex sympathetic dystropy | What patient and injury factors predict outcome following wrist fracture and determine which patients may benefit from the various treatment options? |
| Different methods of external fixation for treating distal radial fractures in adults.<br>Cochrane Database of Systematic Reviews, no. 1 (2008)<br>http://onlinelibrary.wiley.com/doi/10.1002/14651858.CD006522.pub2/abstract | D | Adults with dorsally displaced distal radius fractures | External fixation method 1 (bridging, pins and plaster, extra pin to "floating" fragment, coated or uncoated pins) | External fixation method 2 (bridging, pins and plaster, extra pin to "floating" fragment, coated or uncoated pins) | Functional outcome, re-intervention, Quality of life, pain, complications, cost | In adults with an unstable, dorsally displaced fracture of the distal radius, is [one method of external fixation] superior to [another method] in terms of functional outcome, quality of life, complications, pain, re-intervention or cost? | There is insufficient robust evidence to determine the relative effects of different methods of external fixation. Adequately powered studies could provide better evidence. | What patient and injury factors predict outcome following wrist fracture and determine which patients may benefit from the various treatment options? |
| | | | | | | | | Which hand therapy techniques enable the most efficient return to full function when a condition is treated without an operation, or following surgery or injury? |
| NICE: Rheumatoid arthritis in adults: management : guidance (CG79) | A | Peole with rheumatoid arthritis with pain and/or dysfunction of the hands or wrist | Conservative- stretching and strengthening | | Functional, clinical | | In people with RA what does hand exercise improve functional and clinical outcomes | |
| | | | | | | | | Which hand therapy techniques enable the most efficient return to full function when a condition is treated without an operation, or following surgery or injury? |
| | | In people with OA of the hand | Conservative treatment- splinting | | Functional, clinical | In people with OA what is the benefit in terms of clinical or functional outcomes from the use of splints or braces? | In people with OA which splints or braces are most beneficial and in which subgroups of people with osteoarthritis do they have the greatest benefit? | Which hand therapy techniques enable the most efficient return to full function when a condition is treated without an operation, or following surgery or injury? |
| NICE: Osteoarthritis: care and management : guidance (CG177) | A | | | | | | | |
| | | In people with OA of the hand | Coservative treatment- exercise | | Functional, clinical | In people with OA of the hand what is the benefit in terms of clinical and functional outcome from exercise including strengthening? | In people with OA exercise including muscle strengthening should be offered as a core treatment. | Which hand therapy techniques enable the most efficient return to full function when a condition is treated without an operation, or following surgery or injury? |
| NICE: Stroke rehabilitation in adults : guidance (CG162) | A | In people with hand or upper limb dysfunction following stoke | Conservative treatment- functional electrical stimulation | | Functional | In people with hand or upper limb dysfunction following stroke what is the functional benefit, and at what frequency and dose, from functional electical stimulation? | What is the clinical and cost effectiveness of electrical stimulation (ES) as an adjunct to rehabilitation to improve hand and arm function in people after stroke, from early rehabilitation through to use in the community? What are the dose, practice parameters and rehabilitation programme content needed to effect change in hand and arm function with ES. | |
| SIGN: Guideline 118: Management of patients with stroke: rehabilitation, prevention and management of complications, and discharge planning - Full guideline [PDF] | A | In people with hand or upper limb dysfunction following stoke | Conservative treatment- electrical stimulation, biofeedback, virtual reality training or bilateral training | | Functional | In people with hand or upper limb dysfunction following stroke what is the functional benefit from electical stimulation, biofeedback, virtual reality training or bilateral upper limb training? | There is currently insufficient high quality evidence to support or refute the use of electrostimulation, biofeedback, or virtual reality training or bilateral training for improving upper limb function after stroke. | |
| Assistive technology, including orthotic devices, for the management of contractures in adult stroke patients.<br>Cochrane Database of Systematic Reviews, no. 10 (2013)<br>http://onlinelibrary.wiley.com/doi/10.1002/14651858.CD010779/abstract | B | Adult acute and chronic stroke patients | Conservative treatment- electrical, mechanical and electromechanical devices | no treatment; routine therapy; or alternative assistive technology | Clinical, Functional | In adults with joint contracture secondary to stroke what is the best assistive technology in terms of clinical and functional outcomes? | "Despite the lack of underpinning evidence various types of splinting are recommended, such as splinting in the submaximal stretched position and splinting in the functional position (Milazzo 1998)". "Splinting in the fully stretched position has not been offered by therapists to patients, thereby depriving patients of a potentially effective treatment (Lannin 2011)". "The current literature does not differentiate between stretch used on a muscle crossing one joint and those crossing more than one joint". | |
| | | | Conservative treatment- active & passive exercise, positioning, | | | | | |
| Physical treatment interventions for managing spasticity after stroke.<br>Cochrane Database of Systematic Reviews, no. 7 (2011)<br>http://onlinelibrary.wiley.com/doi/10.1002/14651858.CD009188/abstract | B | Adults with hand or upper limb dysfunction following stroke | | Control, placebo or no intervention | Clinical, functional, Quality of life, complications, resources | In adults who are developing spasticity following stroke what are the benefits in terms of improved function and quality of life and decreased impairement and economic burden from physical treatments including active and passive exercise and positioning as compared with placebo, control or no intervention? | What physical treatment interventions including active exercise; passive exercise/stretching and positioning are effective in preventing or minimising impairment, burden of care, patient quality of life and economic burden in those patients developing spasticity post stroke. | |
| | | | | | | | | Which hand therapy techniques enable the most efficient return to full function when a condition is treated without an operation, or following surgery or injury? |
| Exercise therapy for the rheumatoid hand.<br>Cochrane Database of Systematic Reviews, no. 4 (2012)<br>http://onlinelibrary.wiley.com/doi/10.1002/14651858.CD003832.pub2/abstract | B | In people with rheumatoid arthritis | Conservative treatment- exercise | control exercise therapy | functional, clinical, adverse events, patient satisfaction, economic burden | In people with rheumatoid arthritis what is the effectiveness of hand exercise in improving upper limb function? | It is unclear whether specific exercise for people with rheumatoid arthritis is effective in improving upper limb function. | |
| | | | | | | | | Which hand therapy techniques enable the most efficient return to full function when a condition is treated without an operation, or following surgery or injury? |
| Exercises for hand osteoarthritis.<br>Cochrane Database of Systematic Reviews, no. 2 (2013)<br>http://onlinelibrary.wiley.com/doi/10.1002/14651858.CD010388/abstract | E | In people with hand osteoarthritis | Conservative treatment, exercise | control (no exercise) or other exercise | functional, clinical | In people with hand osteoarthritis what are the beneficial effects of exercise on hand pain, function and finger joint stiffness? | There is low quality evidence with a small effect size demonstrating benefit from hand exercise in people with OA. It is unclear if these effects represent clinically important change. | Which hand therapy techniques enable the most efficient return to full function when a condition is treated without an operation, or following surgery or injury? |
| Hands-on therapy interventions for upper limb motor dysfunction following stroke.<br>Cochrane Database of Systematic Reviews, no. 6 (2011)<br>http://onlinelibrary.wiley.com/doi/10.1002/14651858.CD006609.pub2/abstract | D | People with stroke and impaired upper limb function | Conservative- stretching, passive exercises and mobilization | Control, other therapy | Functional, clinical | In adults with stroke do specific hands-on therapeutic interventions enhance motor activity and function of the upper limb? | Therapists use a variety of techniques to help the arm to get better following stroke. However, the details of what they do are unclear, are not well described in studies, and are used in various combinations. It is not known which elements of these techniques are effective. | |

| | | | | | | | | |
|---|---|---|---|---|---|---|---|---|
| Interventions for improving upper limb function after stroke.<br>Cochrane Database of Systematic Reviews, no. 11 (2014)<br>http://onlinelibrary.wiley.com/doi/10.1002/14651858.CD010820.pub2/abstract | D | Peole with impaired upper limb function following stroke | Conservative- exercise including constraint induced movement therapy, mirror therapy, biofeedback, Bobath therapy, electrical stimulation, reach-to-grasp exercise, repetitive task training, strength training and stretching and positioning | | Functional, clincial | In people who have had a stroke, what interventions help to promote arm and hand recovery | No high level evidence can be found for any interventions that are currently used in routine practice in people who have had a stroke which affects the hand or upper limb. Recommendations for future research include adequately powered, high-quality RCTs to confirm the benefit of CIMT, mental practice, mirror therapy, virtual reality, transcranial magnetic stimulation (rTMS) and up-to-date reviews related to biofeedback, Bobath therapy, electrical stimulation, reach-to-grasp exercise, repetitive task training, strength training and stretching and positioning. | |
| Rehabilitation following carpal tunnel release.<br>Cochrane Database of Systematic Reviews, no. 2 (2016)<br>http://onlinelibrary.wiley.com/doi/10.1002/14651858.CD004158.pub3/abstract | D | In people who have had carpal tunnel surgery | Conservative tx including wrist splinting, dressings, exercise, controlled cold therapy, ice therapy, multi-modal hand rehabilitation, laser therapy, electrical modalities, scar desensitisation, and arnica | Placebo, no intervention or other rehab | Functional, clinical | In people who have undergone carpal tunnel surgery what is the effectiveness and safety of rehabilitation interventions compared with no treatment, placebo, or another intervention? | There is limited low quality evidence for the benefit of the rehab interventions for people who have undergone CTS surgery | Which hand therapy techniques enable the most efficient return to full function when a condition is treated without an operation, or following surgery or injury? |
| Exercise and mobilisation interventions for carpal tunnel syndrome.<br>Cochrane Database of Systematic Reviews, no. 6 (2012)<br>http://onlinelibrary.wiley.com/doi/10.1002/14651858.CD009899/abstract | D | In people with carpal tunnel syndrome | Conservative- exercise | No treatment, placebo, or other non-surgical intervention | Clinical, functional | In people with carpal tunnel syndrome, what is the efficacy and safety of exercise and mobilisation interventions compared with no treatment, a placebo or another non-surgical intervention in people with CTS? | There is limited and very low quality evidence of benefit for exercise and mobilisation interventions for CTS. | Which hand therapy techniques enable the most efficient return to full function when a condition is treated without an operation, or following surgery or injury? |
| Ergonomic positioning or equipment for treating carpal tunnel syndrome.<br>Cochrane Database of Systematic Reviews, no. 1 (2012)<br>http://onlinelibrary.wiley.com/doi/10.1002/14651858.CD009600/abstract | D | In people with carpal tunnel syndrome | Conservative- ergononic equipment or positioning | no treatment, placebo or other non-surgical intervention | Clinical | In people with carpal tunnel syndrome, what are the effects of ergonomic positioning or equipment compared with no treatment, a placebo or another non-surgical intervention in people with CTS | There is insufficient evidence from randomised controlled trials to determine whether ergonomic positioning or equipment is beneficial or harmful for treating carpal tunnel syndrome | Which hand therapy techniques enable the most efficient return to full function when a condition is treated without an operation, or following surgery or injury? |
| Occupational therapy for rheumatoid arthritis.<br>Cochrane Database of Systematic Reviews, no. 1 (2004)<br>http://onlinelibrary.wiley.com/doi/10.1002/14651858.CD003114.pub2/abstract | No uncertainty | In people with rheumatoid arthritis | Conservative- training, education, counseling, splinting | Control | Functional, quality of life | In people with rheumatoid arthritis do OT interventions including training of motor function, training of skills, instruction on joint protection and energy conservation, counseling, instruction about assistive devices and provision of splints improve outcome on functional ability, social participation and/or health related quality of life | There is "gold" level evidence that occupational therapy can help people with rheumatoid arthritis to do daily chores such as dressing, cooking and cleaning and with less pain. Benefits are seen with occupational therapy that includes training, advice and counseling and also with advice on joint protection. Splints can decrease pain and improve grip strength. | |
| Splints and orthosis for treating rheumatoid arthritis.<br>Cochrane Database of Systematic Reviews, no. 4 (2001)<br>http://onlinelibrary.wiley.com/doi/10.1002/14651858.CD004018/abstract | E | In people with rheumatoid arthritis | Conservative- splinting | Control, placebo or other orthosis | clinical, functional | In people with RA, what is the effectiveness of splints/orthoses in relieving pain, decreasing swelling, and/or preventing deformity, and what is the impact of splints/orthoses on strength, mobility, and function? | There is insufficient evidence to make firm conclusions about the effectiveness of working wrist splints in decreasing pain or increasing function for people with RA. Preliminary evidence suggests that resting hand and wrist splints do not seem to affect range of motion (ROM) or pain, although participants prefer wearing a resting splint to not wearing one. | |
| Post-operative therapy for metacarpophalangeal arthroplasty.<br>Cochrane Database of Systematic Reviews, no. 1 (2008)<br>http://onlinelibrary.wiley.com/doi/10.1002/14651858.CD003522.pub2/abstract | D | In people with rheumatoid arthritis | Conservative- exercise, splinting | Control | Clinical, functional | In people with rheumatoid arthritis what is the effectiveness of post-operative therapy regimes including splinting and exercise for increasing hand function after MCP arthroplasty ? | Well-designed randomised controlled trials which compare the efficacy of different therapeutic splinting programmes following MCP arthroplasty are required. The results of one study (silver level evidence) suggest that continuous passive motion alone is not recommended for increasing motion or strength after MCP arthroplasty | |
| Rehabilitation for distal radial fractures in adults.<br>Cochrane Database of Systematic Reviews, no. 9 (2015)<br>http://onlinelibrary.wiley.com/doi/10.1002/14651858.CD003324.pub3/abstract | E | Adults with distal radius fractures | Conservative- exercise | No intervention, control | Clinical, functional | In adults with conservatively or surgically treated distal radial fractures what are the effects of rehab interventions including active and passive range of motion exercise and ADL training? | The available evidence from RCTs is insufficient to establish the relative effectiveness of active and passive range of motion or ADL training in the rehabilitation of adults with fractures of the distal radius. | What factors predict the greatest benefit from intensive hand therapy following injury? |
| Physiotherapy for pain and disability in adults with complex regional pain syndrome (CRPS) types I and II.<br>Cochrane Database of Systematic Reviews, no. 2 (2016)<br>http://onlinelibrary.wiley.com/doi/10.1002/14651858.CD010853.pub2/abstract | E | Adults with CRPS types 1 and II | Conservative- exercise, education, manual therapy, sensory motor rehab | placebo, no treatment | Clinical, functional | In adults with CPRS types I and II what is the effectiveness of physioptherapy interventions including manual therapy, therapeutic exercise,electrotherapy, education and cortically directed sensory-motor rehabilitation strategies compared with placebo, no treatment in reducing pain and disability? | The best available data show that graded motor imagery and mirror therapy may provide clinically meaningful improvements in pain and function in people with CRPS I although the quality of the supporting evidence is very low. Evidence of the effectiveness of multimodal physiotherapy, electrotherapy and manual lymphatic drainage for treating people with CRPS types I and II is generally absent or unclear. Large scale, high quality RCTs are required to test the effectiveness of physiotherapy-based interventions for treating pain and disability of people with CRPS I and II. | |
| Vocational rehabilitation for enhancing return-to-work in workers with traumatic upper limb injuries.<br>Cochrane Database of Systematic Reviews, no. 10 (2013)<br>http://onlinelibrary.wiley.com/doi/10.1002/14651858.CD010002.pub2/abstract | C | Adult workers with traumatic upper limb injuries | Conservative | Alternative control intervention such as standard rehabilitation, an incomplete form of the vocational rehabilitation intervention, or waiting-list controls. | Functional | In adult workers with traumatic upper limb injuries what is the effectiveness of vocational rehabilitation programs in enhancing return to work, decreasing disability, reducing cost burden and improving productivity? | There is currently no high-level evidence to support or refute the efficacy of vocational rehabilitationin enhancing RTW in workers with traumatic upper limb injuries. Further high-quality RCTs assessing the efficacy of vocational rehabilitation for workers with traumatic upper limb injury are needed to fill this gap in knowledge | Which hand therapy techniques enable the most efficient return to full function when a condition is treated without an operation, or following surgery or injury? |

| | | | | | | | | |
|---|---|---|---|---|---|---|---|---|
| | E | | Surgery (Simple decompression) | Surgery (all types of transposition) | Functional, clinical, complications, quality of life | | | |
| Treatment for ulnar neuropathy at the elbow. Cochrane Database of Systematic Reviews, no. 11 (2016) | C | People with clinical symptoms suggesting the presence of ulnar neuropathy at the elbow, with or without neurophysiological evidence of entrapment. | Surgery | Conservative treatments | Functional, clinical, complications, quality of life | In people with clinical symptoms of ulnar neuropathy at the elbow, what is the best surgical or conservative treatment ... in terms of functional outcomes, clinical outcomes, complications and quality of life? | "The available evidence is not sufficient to identify the best treatment based on clinical, neurophysiological, and imaging characteristics." "No evidence is available on the effects of surgery on quality of life and imaging characteristics of the ulnar nerve at the elbow."<br>"Future research in this area should include RCTs to evaluate the effectiveness of conservative treatments."<br>"Moreover, there is a need for a trial comparing conservative and surgical treatment." | What are the most effective non-surgical methods of treating peripheral nerve compression (for example carpal tunnel syndrome or cubital tunnel syndrome)? AND When is surgery of a greater benefit than non-surgical methods in the treatment of peripheral nerve compression (for example carpal or cubital tunnel syndrome)? |
| | D | | Conservative treatment | Conservative treatment | Functional, clinical, complications, quality of life | | | |
| Conservative treatment for closed fifth (small finger) metacarpal neck fractures.<br>Cochrane Database of Systematic Reviews, no. 3 (2005)<br>http://onlinelibrary.wiley.com/doi/10.1002/14651858.CD003210.pub3/abstract<br>Cochrane Database of Systematic Reviews, no. 3 (2005) | D | Adults with closed fifth metacarpal neck fractures | Functional treatment | Immobilization - conservative treatment | Functional, clinical, complications, quality of life | In adults with closed fifth (small finger) metacarpal neck fractures, is functional treatment better than immobilization ... in terms of functional outcomes, clinical outcomes, complications, and quality of life? | "There was no statistically significant difference in range of motion between any functional treatment (functional taping or compression bandage) and immobilization (plaster cast with immobilization of the MCP and wrist joints) at any point in time (one week, three to six weeks or three to six months follow up)."<br>"This review included no study that documented our primary outcome measure of interest, validated hand function. Therefore, no single treatment regimen can be recommended for all participants." | Which hand/finger/thumb injuries would benefit from surgical intervention over hand therapy or no formal treatment, considering both functional outcome and societal cost? |
| | D | | Different periods and types of immobilization | Different periods and types of immobilization | Functional, clinical, complications, quality of life | In adults with closed fifth (small finger) metacarpal neck fractures, what period and type of immobilization is best ...in terms of functional outcomes, clinical outcomes, complications, quality of life? | | Which hand/finger/thumb injuries would benefit from surgical intervention over hand therapy or no formal treatment, considering both functional outcome and societal cost? |
| Conservative interventions for treating hyperextension injuries of the proximal interphalangeal joints of the fingers.<br>Cochrane Database of Systematic Reviews, no. 2 (2013)<br>http://onlinelibrary.wiley.com/doi/10.1002/14651858.CD009030.pub2/abstract<br>Cochrane Database of Systematic Reviews, no. 2 (2013) | D | People with acute hyperextension injuries of the proximal interphalangeal joints of the fingers managed conservatively (without surgery) ~~non-surgically~~ | Conservative treatment (unrestricted movement, buddy strapping, protective splinting, and immobilisation) | Conservative treatment (unrestricted movement, buddy strapping, protective splinting, and immobilisation) | Functional, clinical, quality of life | In people with acute hyperextension injuries of the proximal interphalangeal joints of the fingers, which type of conservative treatment is best ... in terms of functional, clinical outcomes, quality of life? | "The quality of evidence available for the purpose of this review is very low."<br>"There is insufficient evidence from trials testing the need for, and the extent and duration of, immobilisation to inform on the key conservative management decisions for treating hyperextension injuries of the proximal interphalangeal joints." | |
| | | | Different timeframes of conservative treatment | Different timeframes of conservative treatment | Functional, clinical, quality of life | | | |
| | D | | Conservative treatments | Conservative treatments, including no intervention | Functional, clinical, complications~~,~~ resources, patient satisfaction/adherence | | | |
| Interventions for treating mallet finger injuries.<br>Cochrane Database of Systematic Reviews, no. 3 (2004)<br>http://onlinelibrary.wiley.com/doi/10.1002/14651858.CD004574.pub2/abstract<br>Cochrane Database of Systematic Reviews, no. 3 (2004) | D | Patients of any age with mallet finger injury or deformity ~~People with mallet finger injuries~~ | Surgical treatments | Conservative treatments, including no intervention | Functional, clinical, complications~~,~~ resources, patient satisfaction/adherence | In people with mallet finger injuries, what is the best surgical or conservative treatment ... in terms of functional outcomes, clinical outcomes, complications, resource use and patient satisfaction/adherence? | "There was insufficient evidence from comparisons tested within randomised controlled trials to establish the relative effectiveness of different, either custom-made or off-the-shelf, finger splints used for treating mallet finger injury. There was a useful reminder that splints used for prolonged immobilisation should be robust enough for everyday use, and of the central importance of patient adherence to instructions for splint use. There was insufficient evidence to determine when surgery is indicated." | |
| | D | | Surgical treatments | Surgical treatments | Functional, clinical, complications~~,~~ resources, patient satisfaction/adherence | | | |
| Antibiotics for preventing infection in open limb fractures.<br>Cochrane Database of Systematic Reviews, no. 1 (2004)<br>http://onlinelibrary.wiley.com/doi/10.1002/14651858.CD003764.pub2/abstract<br>Cochrane Database of Systematic Reviews, no. 1 (2004) | E | People of any age with open fractures of the limbs, including people with open finger fractures | Antibiotic administered before or at the time of primary treatment of the open fracture | Placebo or no antibiotic | Early wound infection, other complications, clinical | In people with open finger fractures, what is the effectiveness of antibiotic prophylaxis ... in terms of reduced early wound infections, other complications, clinical outcomes and resource use? | "The use of antibiotics had a protective effect against early infection compared with no antibiotics or placebo (risk ratio 0.43 (95% confidence interval (CI) 0.29 to 0.65); absolute risk reduction 0.07 (95% CI 0.03 to 0.10)."<br>"Subgroup analysis (Analysis 1.3) explored the importance of fracture location and found that in open finger fractures, there was no evidence of significant benefit from antibiotics (3 trials, 367 participants; RR 0.56, 95% CI 0.26 to 1.23). In trials which did not include open finger fractures, antibiotics significantly reduced early infection (4 trials, 472 participants, RR 0.37, 95% CI 0.21 to 0.68)."<br>"Further placebo controlled trials to evaluate the effectiveness of antibiotics for ooen fractures of the limbs proximal to | Does the use of antibiotics affect the infection rate following hand trauma or elective hand surgery? |
| Percutaneous pinning for treating distal radial fractures in adults.<br>Cochrane Database of Systematic Reviews, no. 3 (2007)<br>http://onlinelibrary.wiley.com/doi/10.1002/14651858.CD006080.pub2/abstract<br>Cochrane Database of Systematic Reviews, no. 3 (2007) | E | Adults with a fracture of the distal radius | Percutaneous pinning | Conservative interventions, such as plaster cast immobilisation | Functional, clinical, complications, resource use | In adults with fractures of the distal radius, is percutaneous pinning better than conservative treatment, such as plastercast immobilisation, ... in terms of functional outcomes, clinical outcomes, complications and resource use? | established. The higher rates of complications with Kapandji pinning and biodegradable materials casts some doubt on their general use."<br>"Only a few and provisional conclusions relating to clinical management can be drawn from the available randomised trials.<br>For dorsally displaced fractures, across-fracture percutaneous pinning helps to maintain reduced positions and thereby reduce deformity and malunion compared with plaster cast immobilisation alone. There is limited evidence that its use improves function. Complications are usually minor and, to some extent, avoidable. However, uncertainty remains about the indications for percutaneous pinning, the best technique to employ, and the extent and duration of immobilisation. Kapandji pinning, involving the support rather than fixation of the distal fracture fragment, appears to be associated with a less favourable outcome, particularly an excess of probable iatrogenic complications. More recent refinements of this technique have yet to be evaluated. There was some evidence of an excess of complications which, coupled with the extra demands at surgery, are likely to outweigh the putative advantages - avoidance of metal wire extraction and the associated risk of damage - of biodegradable pins." | |
| | D | | Different methods of percutaneous pinning | Different methods of percutaneous pinning | Functional, clinical, complications, resource use | In adults with fractures of the distal radius, which method of percutaneous pinning is best ... in terms of functional outcomes, clinical outcomes, complications and resource use? | | |
| | E | | Cleansing of pin sites | No cleansing of pin sites | Incidence of infection, other complications, clinical, patient acceptability and resource use. | | | |
| | E | | Different cleansing solutions for pin site care | Different cleansing solutions for pin site care | Incidence of infection, other complications, clinical, patient acceptability and resource use. | | | |
| | E | | Different methods of cleansing for pin site care | Different methods of cleansing for pin site care | Incidence of infection, other complications, clinical, patient acceptability and resource use. | | | |
| Pin site care for preventing infections associated with external bone fixators and pins.<br>Cochrane Database of Systematic Reviews, no. 12 (2013)<br>http://onlinelibrary.wiley.com/doi/10.1002/14651858.CD004551.pub3/abstract<br>Cochrane Database of Systematic Reviews, no. 12 (2013) | E | Adults and children with pins inserted for either external fixators or skeletal traction | Dressing for pin site care | No dressing for pin site care | Incidence of infection, other complications, clinical, patient acceptability and resource use. | In adults and children with pins inserted for either external fixators or skeletal traction orthopaedic percutaneous pin sites, what is the effectiveness of cleansing, massage and dressing techniques for the prevention of infections...in terms of the incidence of infection, complications, clinical outcomes, patient acceptability and resource use? | "In conclusion, this review has not found sufficient evidence to recommend a particular strategy of pin site care. Adequately-powered, randomised trials are required to examine the effects of different pin care regimens and their co-interventions such as antibiotic use; furthermore, other extraneous factors must be controlled in the study designs."<br>"It is not surprising that few differences were reported between pin site care regimens in the included studies, as few studies ensured that the sample size was adequately powered to find differences."<br>"Overall, study quality was poor. Although randomised, most studies were small and underpowered with methodological flaws, and only two were blinded. In most studies, minimal efforts were made to control for other factors that might have influenced the results, such as patient compliance with pin care protocols after discharge from hospital, antibiotic use, length of time pins were in place, location of pins, pin insertion procedures, types of hardware used, heterogeneity (variation) in the age of participants and condition being treated by external fixators." | |
| | D | | Different types of dressing for pin site care | Different types of dressing for pin site care | Incidence of infection, other complications, clinical, patient acceptability and resource use. | | | |
| | C | | Massage for pin site care | No massage for pin site care | Incidence of infection, other complications, clinical, patient acceptability and resource use. | | | |
| | C | | Different methods of massage | Different methods of massage | Incidence of infection, other complications, clinical, patient acceptability and resource use. | | | |
| | | | Mechanical reduction using finger trap traction | Manual traction | Succesful reduction, functional, clinical, resource use | | | |
| | | | | | | | "There was insufficient evidence from comparisons tested within randomised controlled trials to establish the relative | |

| | | | | | | | | |
|---|---|---|---|---|---|---|---|---|
| Closed reduction methods for treating distal radial fractures in adults.Closed reduction methods for treating distal radial fractures in adults.<br>Cochrane Database of Systematic Reviews, no. 1 (2003)<br>http://onlinelibrary.wiley.com/doi/10.1002/14651858.CD003763/abstract | D | Skeletally mature adults with a displaced and closed fracture of the distal radius considered suitable for reduction | Manual reduction with counter-traction provided by patient | Manual reduction with counter-traction provided by an assistant | Succesful reduction, functional, clinical, resource use | In adults with a displaced and closed fracture of the distal radius, what is the relative effectiveness of different conservative (non-surgical) methods used for closed reduction … in terms of succesful reduction, functional and clinical outcomes and resource use? | [illegible] effectiveness of different methods of closed reduction used in the treatment of displaced fractures of the distal radius in adults."<br>"Research on closed reduction methods needs to be set in the context of the overall management of these fractures. There are many unresolved issues such as:<br>•When is reduction required and what is the best method of anaesthesia, if such is necessary?<br>•When, to what extent, by what means and for how long is immobilisation required?<br>•When is surgery indicated and what method should be used?" | |
| | | | Mechanical reduction using finger traps and a dynamic device | Manual reduction | Succesful reduction, functional, clinical, resource use | | | |
| | D | | Implantation of bone grafts or substitutes alone | Conservative interventions such as plaster cast immobilisation | Functional, clinical, complications, resource use | | | |
| | D | | Implantation of bone grafts or substitutes along with surgical fixation (percutaneous pinning, external fixation, internal fixation or combinations of these) | The same method of surgical fixation alone | Functional, clinical, complications, resource use | | | |
| Bone grafts and bone substitutes for treating distal radial fractures in adults.<br>Cochrane Database of Systematic Reviews, no. 2 (2008)<br>http://onlinelibrary.wiley.com/doi/10.1002/14651858.CD006836.pub2/abstract<br>Cochrane Database of Systematic Reviews, no. 2 (2008) | D | Skeletally mature adults with distal radius fracture | Implantation of bone grafts or substitutes alone | Surgical fixation (percutaneous pinning, external fixation, or combinations of these). | Functional, clinical, complications, resource use | In adults with a distal radius fracture, what is the effectiveness of using bone scaffolding materials (bone grafts and substitutes) … in terms of functional outcomes, clinical outcomes, complications and resource use? | "There is some evidence that bone scaffolding may improve anatomical outcome compared with plaster cast immobilisation alone but there is insufficient evidence on functional outcome and safety. There is insufficient evidence on the effectiveness of bone scaffolding supplementary to external fixation, or relative to percutaneous pinning or to external fixation; or of different methods of bone scaffolding." | |
| | D | | Different types of bone scaffolding (e.g. autografts versus allografts; grafts versus bone substitutes; bioabsorbable versus bio-inert substitute materials). | Different types of bone scaffolding (e.g. autografts versus allografts; grafts versus bone substitutes; bioabsorbable versus bio-inert substitute materials). | Functional, clinical, complications, resource use | | | |
| | C | | Different types and durations of immobilisation after bone scaffolding | Different types and durations of immobilisation after bone scaffolding | Functional, clinical, complications, resource use | | | |
| Anaesthesia for treating distal radial fracture in adults.<br>Cochrane Database of Systematic Reviews, no. 3 (2002)<br>http://onlinelibrary.wiley.com/doi/10.1002/14651858.CD003320/abstract<br>Cochrane Database of Systematic Reviews, no. 3 (2002) | E | Skeletally mature adults of either sex who had completed skeletal growth and who were receiving conservative or surgical treatment for a fracture of the distal radius. | Different types and physical techniques of anaesthesia used for treating distal radial fractures (too many comparisons to list here) | Different types and physical techniques of anaesthesia used for treating distal radial fractures (too many comparisons to list here) | Successful anaesthesia, complications, functional, clinical outcomes and resource use | In adults receiving conservative or surgical treatment for a fracture of the distal radius, what is the relative effectiveness of different types and physical techniques of anaesthesia … in terms of successful anaesthesia, complications, adverse effects, functional, clinical outcomes and resource use ? | "There was insufficient robust evidence from randomised trials to establish the relative effectiveness of different methods of anaesthesia, different associated physical techniques or the use of drug adjuncts in the treatment of distal radial fractures. There is, however, some indication that haematoma block provides poorer analgesia than IVRA, and can compromise reduction." | |
| Interventions for treating pain and disability in adults with complex regional pain syndrome- an overview of systematic reviews.<br>Cochrane Database of Systematic Reviews, no. 4 (2013)<br>http://onlinelibrary.wiley.com/doi/10.1002/14651858.CD009416.pub2/abstract<br>Cochrane Database of Systematic Reviews, no. 4 (2013) | E | Adults described as suffering from CRPS or an alternative descriptor for this condition (for example reflex sympathetic dystrophy, causalgia) | Any ~~t~~ intervention~~s~~ aimed at reducing pain, disability, or both | Any intervention~~s~~ aimed at reducing pain, disability, or both | Functional (pain & disability), complications, quality of life, patient satisfaction | In adults described as suffering from complex regional pain syndrome, what is the relative effectiveness of different interventions to reduce pain and disability...in terms of ...? | [illegible] However, there is moderate quality evidence that IVRB guanethidine is not effective. "There is low or very low quality evidence relating to the efficacy of a range of therapies in CRPS although all of this evidence, both positive and negative, should be interpreted with caution and does not reliably aid clinical decision making. Until further larger trials are undertaken an evidence-based approach to managing CRPS will remain difficult."<br>"There is a clear need for further research for most existing treatment for CRPS as reasonably confident conclusions can only be drawn for the ineffectiveness of IVRB guanethidine." | Can hand therapy or hand surgery help in the management of complex regional pain syndrome? |
| NICE: Fractures (non-complex): assessment and management : guidance (NG38) | A | Patients with displaced distal radius fractures | Manipulation with real -time image guidance | Manipulation without real -time image guidance | Clinical, Functional (manipulation and reduction of bone fragments into correct anatomical position) | For patients with displaced fractures of the distal radius, is manipulation with real -time image guidance more clinically and cost effective than manipulation without real -time image guidance? | "In a large minority of patients with a distal radius fracture, the bone fragments are displaced and need manipulation and reduction into an anatomical position. Currently in the NHS, most manipulations for distal radius fractures are performed in the emergency department without real -time image guidance. It is believed that image guidance may be important, but despite hundreds of people having manipulation for these fractures in the emergency department each day, there are no high-quality studies in this area." | What is the best treatment of wrist fractures regarding patient outcomes and cost? |
| | | Children, young people and adults that require surgery following a distal radial fracture, after experiencing a traumatic incident | ≥14 days post injury, 8–13 days post injury, >48 hours to ≤7 days post injury, Within 48 hours | Comparison of the above | Critical: ⍰ Health-related quality of life ⍰ Need for re-operation ⍰ PROMS ⍰ Wound infection ⍰ Anaesthetic complications ⍰ Growth plate arrest Important: ⍰ Pain/discomfort ⍰ Return to normal activities ⍰ Psychological wellbeing | Review question: What is the maximum safe delay in surgical management of fractures of the distal radius before outcome is compromised? | Pg 185-187 full guideline document. "No RCTs or cohort studies were found for this review question." "No published evidence was available so recommendations were made by consensus." "Despite the lack of clinical evidence, the GDG felt that this was too urgent an issue for a research recommendation. It was felt that at present many intra-articular distal radius surgeries are carried out too late leading to possibly poorer outcomes. Such delays were usually made for non-clinical reasons. It was therefore felt that a clinical recommendation was needed to encourage a change in practice. The time frames suggested are based upon clinical experience, knowledge of physiological healing times, and what is achievable within the NHS." | What is the best treatment of wrist fractures regarding patient outcomes and cost? |
| | | Children, young people and adults experiencing a dorsally displaced fracture of the distal radius (without neurovascular compromise) | Closed reduction and plaster cast immobilisation, Closed reduction and external fixation , Closed reduction and percutaneous wiring , Open reduction and internal fixation (ORIF) , No treatment | Comparison of the above | Critical: ⍰ Health-related quality of life ⍰ Pain/discomfort ⍰ Return to normal activities ⍰ Psychological wellbeing ⍰ Hand and wrist function ⍰ Adverse effects o Pin-site infection o Post traumatic osteoarthritis o Complex regional pain syndrome Important: ⍰ Need for revision surgery ⍰ Need for further surgery (for example, removal of metalwork) ⍰ Number of attendances/bed days ⍰ Radiological anatomical measures | Review question: What is the most clinically and cost effective definitive treatment for dorsally displaced low-energy fractures of the distal radius? | p 188-209 full guideline document "Clinical evidence: The vast majority of the data was at low or very low GRADE quality. Several analyses also demonstrated some unexplained heterogeneity. No subgroup analyses were conducted due to too few studies reporting data separately for the specified subgroups (age and location of fracture)." | What is the best treatment of wrist fractures regarding patient outcomes and cost? |
| | | | | | | Which dorsally displaced distal radial fractures benefit from surgical rather than conservative treatment? | p 209 full guideline "The GDG felt that there is no clear evidence in the literature concerning which dorsally displaced distal radial fractures benefit from surgical rather than conservative treatment." " The GDG considered making a research recommendation in this area. However this was not done because a review question had not been posed on this specific topic." | Which factors indicate who can be treated without an operation rather than with surgery following a wrist fracture? |
| | | Adults with a dorsally displaced distal radius fracture (without neurovascular compromise) due to a traumatic incident | Conscious sedation ⍰ Entonox ⍰ Haematoma block ⍰ IVRA ⍰ Regional nerve block (including brachial plexus block) ⍰ Haematoma block with conscious sedation ⍰ Haematoma block with Entonox | Compared with each other (between categories only) | Critical: ⍰ Health-related quality of life ⍰ Pain ⍰ Need for re-manipulation ⍰ Need for surgical fixation ⍰ Patient-reported function PRWE, DASH ⍰ Death ⍰ Laryngospasm/Respiratory depression ⍰ Nausea/vomiting ⍰ Cardiac arrhythmias ⍰ Nerve damage ⍰ Infection ⍰ Hallucinations/emergent phenomena Important: ⍰ Return to normal activities | Review question: What type of anaesthetic is the most clinically and cost effective for closed reduction of dorsally displaced distal radius fractures in people without neurovascular compromise in the emergency department? | "All but one outcome were graded as low or very low quality evidence. This was due to risk or bias and/or imprecision. Risk of bias was very serious for most outcomes due to a lack of allocation concealment, or a lack of patient, health-care practitioner and assessor blinding. There was serious or very serious imprecision for most outcomes due to the 95% confidence intervals crossing one or both MIDs." "No economic evidence was identified for this question." | What is the best treatment of wrist fractures regarding patient outcomes and cost? |
| | | Children, young people and adults with a suspected scaphoid fracture following a traumatic incident. | ⍰ CT ⍰ MRI ⍰ X-ray | Compared with each other | Critical: ⍰ Time in plaster cast ⍰ Number of outpatient visits ⍰ health-related quality of life ⍰ Pain/discomfort ⍰ Return to normal activities ⍰ Psychological wellbeing ⍰ missed injury ⍰ non-union/malunion ⍰ avascular necrosis ⍰ post-traumatic arthritis ⍰ additional radiation exposure Important: ⍰ Grip strength ⍰ Range of motion | Review question: What is the most clinically and cost-effective imaging strategy for patients with clinically suspected scaphoid fracture? | While diagnostic cohort studies can tell us about the relative accuracy of a diagnostic test compared to a reference standard, they do not tell us whether adopting a particular diagnostic strategy improves patient outcomes. Evidence on patient outcomes is only available from diagnostic randomised controlled trials which compare two diagnostic interventions with identical subsequent treatment as indicated by the diagnostic test. One diagnostic RCT was included, but because this evidence did not cover all the tests in the protocol, diagnostic accuracy studies were also included | |
| NICE: Fractures (complex): assessment and management : guidance (NG37) | No hand relevant research recommend-ations included | | | | | | | NA |
| NICE: Low-intensity pulsed ultrasound to promote fracture healing - guidance (IPG374) | No gap identified and no research recommend-ations made | | | | | | Fractures: non complex | NA |
| NICE: Direct skeletal fixation of limb or digit prostheses using intraosseous transcutaneous implants - guidance (IPG270) | A | Patients who require a prosthesis following upper limb or digit amputation or congenital deficiency | Direct skeletal fixation of upper limb or digit prostheses using an intraosseous transcutaneous implant | Other methods of locating prostheses | Not stated | In patients who require a prosthesis following upper limb or digit amputation or congenital deficiency, what is the safety and efficacy of direct skeletal fixation using intraosseous implants, compared to other methods of location? | Ongoing orthopaedic management | Rare out of scope |
| Peri-operative antibiotics for hand trauma involving tendons and nerves.<br>Cochrane Database of Systematic Reviews, no. 6 (2012)<br>http://onlinelibrary.wiley.com/doi/10.1002/14651858.CD002107.pub2/abstract<br>Cochrane Database of Systematic Reviews, no. 6 (2012) | B | People with traumatic hand wounds involving tendons or a nerve (bone injuries, burns and simple cuts excluded) | Peri-operative antibiotics | No peri-operative antibiotics | Infection, Complications, Clinical, Functional, Resource use | In people with traumatic hand wounds trauma involving tendons or a nerve, what is the effectiveness of peri-operative antibiotic use in reducing infection, other complications and improving clinical and functional outcome? | N/A | Does the use of antibiotics affect the infection rate following hand trauma or elective hand surgery? |
| | | | Peri-operative antibiotic regimes | Other peri-operative antibiotic regimes | | | | |
| | | | Peri-operative antibiotics | Other antimicrobial or anti-infective interventions | | | | |

| | | | | | | | | |
|---|---|---|---|---|---|---|---|---|
| Internal fixation for treating distal radius fractures in adults.<br>Cochrane Database of Systematic Reviews, no. 7 (2014)<br>http://onlinelibrary.wiley.com/doi/10.1002/14651858.CD011213/abstract<br>Cochrane Database of Systematic Reviews, no. 7 (2014) | B | Adults with distal radius fractures | Internal fixation | Conservative treatment (closed reduction and cast immobilisation) | Complications, Clinical, Functional, Quality of life, Patient satisfaction, Resource use | In adults with distal radius fractures, what is the effectiveness of internal fixation compared to conservative treatment (consisting of closed reduction and cast immobilisation)? | N/A | What is the best treatment of wrist fractures regarding patient outcomes and cost? |
| | | | Different types of internal fixation | Different types of internal fixation | Complications, Clinical, Functional, Quality of life, Patient satisfaction, Resource use | In adults with distal radius fractures, what is the comparative effectivenessof different methods of internal fixation? | N/A | What is the best treatment of wrist fractures regarding patient outcomes and cost? |
| | | | Different types and durations of immobilisation after internal fixation | Different types and durations of immobilisation after internal fixation | Complications, Clinical, Functional, Quality of life, Patient satisfaction, Resource use | In adults with distal radius fractures, what is the optimum immobilisation regime after internal fixation? | N/A | What is the best treatment of wrist fractures regarding patient outcomes and cost? |
| Internal fixation versus other surgical methods for treating distal radius fractures in adults.<br>Cochrane Database of Systematic Reviews, no. 7 (2014)<br>http://onlinelibrary.wiley.com/doi/10.1002/14651858.CD011212/abstract<br>Cochrane Database of Systematic Reviews, no. 7 (2014) | B | Adults with distal radius fractures | Internal fixation | Other methods of surgery that are not internal fixation (percutaneous pinning or external fixation ) | Complications, Clinical, Functional, Quality of life, Patient satisfaction, Resource use | In adults with distal radius fractures, what is the effectiveness of internal fixation compared to other methods of surgery (percutaneous pinning or external fixation)? | N/A | What is the best treatment of wrist fractures regarding patient outcomes and cost? |
| Interventions for treating ulnar collateral ligament injuries of the thumb.<br>Cochrane Database of Systematic Reviews, no. 8 (2014)<br>http://onlinelibrary.wiley.com/doi/10.1002/14651858.CD011267/abstract<br>Cochrane Database of Systematic Reviews, no. 8 (2014 | B | People with an acute or chronic ulnar collateral ligament injury of the thumb | Different methods of surgical or non-surgical (conservative) treatment | Different methods of surgical or non-surgical (conservative) treatment | Clinical, Functional, Complications, Resource use | In people with an acute or chronic ulnar collateral ligament injury of the thumb, what is the most effective surgical or non-surgical (conservative) intervention? | N/A | Which hand/finger/thumb injuries would benefit from surgical intervention over hand therapy or no formal treatment, considering both functional outcome and societal cost? |
