## Supplemental file 4 for "Uncertainty extraction and verification for the Common Conditions Affecting the Hand and Wrist Priority Setting Partnership: A proposed methodology for Priority Setting Partnerships with the James Lind Alliance"

**Uncertainty long list:**

| | Uncertainty | Reason |
|---|---|---|
| 1 | Can a more holistic approach (psychologically, socially, spiritually, culturally informed) hand therapy have additional benefits for patients with common hand conditions including traumatic injuries? | A |
| 2 | Can alternatives to plaster casts be used in the conservative treatment of wrist fractures? | C/D |
| 3 | Can hand therapy or hand surgery help in the management of cold intolerance following hand surgery or injury? | C |
| 4 | Can hand therapy or hand surgery help in the management of complex regional pain syndrome? | C |
| 5 | Can novel approaches to the delivery of hand surgery demonstrate clinical and cost benefits? | A |
| 6 | Can scar formation be manipulated to improve results following hand surgery/trauma? | D |
| 7 | Compared with removing the trapezium bone (a bone at the bottom of the thumb) in arthritis of the base of the thumb, are any other surgical or non-surgical techniques better in terms of patient outcome, power, movement and pain relief? | D |
| 8 | Which hand/finger/thumb injuries would benefit from surgical intervention over hand therapy or no formal treatment, considering both functional outcome and societal cost? | B |
| 9 | Do pre-existing systemic pain conditions (where patients have difficult to treat pain as well as a hand injury or problem) have a negative effect on the outcomes following hand injury or surgery? | B |
| 10 | Does prosthetic (artificial) replacement or hand transplantation have the greater clinical and cost benefits when used in the hand and wrist? | C |
| 11 | Does regional anaesthesia (where the arm is numbed completely) offer benefits directly to the hand and wrist, in the surgical treatment of common hand conditions? | A |
| 12 | Does surgery have a role in the treatment of the complications of Raynaud's disease/systemic sclerosis/scleroderma? | C |
| 13 | Do surgical or non-surgical methods give the best results in post-traumatic stiffness of the proximal interphalangeal joint (the middle knuckle of the finger)? | B |
| 14 | Does the pattern of arthritis in the wrist predict which treatment will give most benefit? | C |

| | Uncertainty | Reason |
|---|---|---|
| 15 | Does the type of metal used (for implants/replacements/plates/screws/wires) during surgery, affect the results? | A |
| 16 | Does the use of antibiotics affect the infection rate following hand trauma or elective hand surgery? | D |
| 17 | Following unsuccessful treatment for an acute tendon injury (snapping or cut), which techniques are most successful at restoring function? | D |
| 18 | How can patient participation in hand therapy be maximised? | A |
| 19 | How is failed joint replacement in the hand/wrist best managed? | A |
| 20 | How is tissue loss in the hand/wrist best treated? | A |
| 21 | In common hand conditions that require splinting to maintain function or prevent deformity, what are the best splint designs and regimens for their use? | C/A |
| 22 | In inflammatory arthropathy (for example rheumatoid arthritis), when should surgical interventions be considered over non-surgical treatments? | A |
| 23 | In patients with Dupuytren's disease, what invasive techniques give the best results in terms of function, recurrence and cost? | C |
| 24 | In patients with ulnar sided wrist pain (pain on the little finger side of the wrist), what interventions give a reliable improvement in function and pain? | B |
| 25 | In previously treated Dupuytren's disease, do any additional techniques (for example splinting, therapy, stretching, massage) help prevent recurrence? | D |
| 26 | In the treatment of common hand conditions, such as peripheral nerve compression syndromes (for example carpal tunnel syndrome), ganglia or arthritis of the fingers/thumb/wrist, do surgical interventions have a demonstrable benefit in patient reported outcome when compared with non-surgical methods or placebo (sham) surgery? | C |
| 27 | In the treatment of broken bones that are not healing in the wrist and hand, which techniques demonstrate the greatest clinical and cost benefits? | B |
| 28 | In the treatment of non-traumatic tendon problems (for example trigger fingers, tendinitis) in the hand and wrist, how does surgical intervention compare to non-surgical methods? | A |

| | Uncertainty | Reason |
|---|---|---|
| 29 | Is denervation (where the nerves that take pain from the joint to the brain are intentionally cut) of a painful joint a better option than alternative surgical treatment, and if so what are the best techniques for denervation and in which joints? | A |
| 30 | Is revision surgery useful in recurrent Dupuytren's disease? | A |
| 31 | Regarding patient and cost benefits, are movement preserving surgeries, such as joint replacement or cartilage replacement, preferential to fusion (permanent stiffening) in the treatment of painful joints in the hand/wrist? | C |
| 32 | What are the best methods of treating tendon injuries? | D |
| 33 | What are the most beneficial interventions in the short and longer term following a burn to the hand/wrist? | A/B |
| 34 | What are the most effective non-surgical methods for treating early arthritis in the hand and fingers? | D |
| 35 | What are the most effective non-surgical methods of treating peripheral nerve compression (for example carpal tunnel syndrome or cubital tunnel syndrome)? | A |
| 36 | What are the most reliable treatments for repetitive strain injury? | C |
| 37 | What factors predict the greatest benefit from hand therapy following injury? | A |
| 38 | What interventions/treatments will have the most positive effect following nerve injury? | B/C |
| 39 | What is the best rehabilitation following treatment for acute or chronic wrist instability? | A |
| 40 | What is the best treatment of wrist fractures regarding patient outcomes and cost? | D |
| 41 | What is the role of steroid injections in the treatment of arthritis of the hand and wrist? | D |
| 42 | What is the role of surgery in Kienbock's disease? | A |
| 43 | What methods are most accurate, user friendly and demonstrate the best clinical utility in measuring patient reported outcomes in common hand conditions? | B |

| | Uncertainty | Reason |
|---|---|---|
| 44 | What non-surgical treatments have benefits over surgery in the treatment of Dupuytren's disease? | B/C |
| 45 | What patient and injury factors predict outcome following wrist fracture and determine which patients may benefit from the various treatment options? | C |
| 46 | What patient or surgical factors may contribute to complications or ongoing symptoms following treatment for common hand conditions? | C |
| 47 | Which surgical procedures (such as finger/hand preserving surgery or amputation) give the best results for cancers of the hand and wrist? | C |
| 48 | What treatments are most effective for the treatment of ongoing symptoms following surgery for carpal or cubital tunnel syndrome (or other entrapped nerves in the arm)? | B |
| 49 | When and how should mucoid cysts/Heberden's nodes be treated? | A/B |
| 50 | When and in whom should treatment for Dupuytren's be commenced? | B |
| 51 | When is surgery of a greater benefit than non-surgical methods in the treatment of peripheral nerve compression (for example carpal or cubital tunnel syndrome)? | B/C |
| 52 | When is the best time to operate following hand injury (for example following fracture, tendon injury or simple skin cuts)? | B |
| 53 | When should patients with pain or deformity following joint damage or arthritis undergo surgery to correct/improve it? | B |
| 54 | Which arthroscopic (keyhole) procedures in the hand/wrist give additional benefits to other surgical techniques or to no intervention? | C |
| 55 | Which factors indicate who can be treated without an operation rather than with surgery following a wrist fracture? | C |
| 56 | Which hand therapy techniques enable the most efficient return to full function following surgery or injury? | C/A |
| 57 | Which patients with a recent scaphoid fracture would benefit from surgery rather than cast or splint treatment? | C |
| 58 | Which patients with acute ligament injuries to the wrist or chronic wrist/distal radio-ulnar joint (the joint on the little finger side of the wrist) instability benefit from surgical treatment rather than from non-surgical methods? | A/B |

| | Uncertainty | Reason |
|---|---|---|
| 59 | Which patients with paralysis, spasticity or functional loss in the upper limb following brain or nerve injury, benefit from surgery in addition to non-surgical treatments? | C |
| 60 | Does the surgical method (i.e. the exact technique used) influence the outcome following surgery for peripheral nerve compression (for example carpal tunnel syndrome or cubital tunnel syndrome)? | C/D |
| 61 | When should trigger digits be referred for specialist treatment? | A |
| 62 | What additions to routine surgical treatment of amputated fingers/hands/arms can help with the survival of the amputated part? | C |
| 63 | Is surgery better than alternative treatments in the management of gouty tophi in the hand and/or wrist? | A |
| 64 | Is local anaesthetic mixed with adrenaline a safe and effective alternative to plain local anaesthetic for use in the hand and digits? | A |
| 65 | What is the best method of detecting the occult or suspected scaphoid fracture? | A |
| 66 | In adults and children with pins inserted for either fixation or skeletal traction (percutaneous pin sites), what is the effectiveness of cleansing, massage and dressing techniques for the prevention of infections, in terms of the incidence of infection, complications, clinical outcomes, patient acceptability and resource use? | D |
| 67 | What are the clinical and cost benefits of non-surgical treatment of spasticity of the upper limb compared to more invasive treatments? | A |
